# Magnetofluidic platform for rapid multiplexed screening of SARS-CoV-2 variants and respiratory pathogens

**DOI:** 10.1101/2021.05.10.21256995

**Authors:** Alexander Y. Trick, Fan-En Chen, Liben Chen, Pei-Wei Lee, Alexander C. Hasnain, Heba H. Mostafa, Karen C. Carroll, Tza-Huei Wang

**Author notes:** Co-first author (Alexander Y. Trick, Fan-En Chen).

## Abstract

The rise of highly transmissible SARS-CoV-2 variants brings new challenges and concerns with vaccine efficacy, diagnostic sensitivity, and public health responses in the fight to end the pandemic. Widespread detection of variant strains will be critical to inform policy decisions to mitigate further spread, and post-pandemic multiplexed screening of respiratory viruses will be necessary to properly manage patients presenting with similar respiratory symptoms. In this work, we have developed a portable, magnetofluidic cartridge platform for automated PCR testing in <30 min. Cartridges were designed for multiplexed detection of SARS-CoV-2 with either distinctive variant mutations or with Influenza A and B. The platform demonstrated a limit of detection down to 2 copies/µL SARS-CoV-2 RNA with successful identification of B.1.1.7 and B.1.351 variants. The multiplexed SARS-CoV-2/Flu assay was validated using archived clinical nasopharyngeal swab eluates (*n* = 116) with an overall sensitivity/specificity of 98.1%/95.2%, 85.7%/100%, 100%/98.2%, respectively, for SARS-CoV-2, Influenza A, and Influenza B. Further testing with saliva (*n* = 14) demonstrated successful detection of all SARS-CoV-2 positive samples with no false-positives.

## Introduction

While the development and distribution of vaccines brings hope for a return to normalcy, extensive diagnostic testing remains critical to curbing the spread of the SARS-CoV-2 virus^1–3^. Even with vaccines, the rise of highly transmissible virus variants that have dominated recent coronavirus disease 2019 (COVID-19) outbreaks raises concerns about mutations leading to potential escape from vaccine protection^4–6^. Insufficient screening and surveillance has left public health officials with large gaps in knowledge of the extent and impact of these variants^7^. Furthermore, demand for testing will continue after the pandemic wanes, as it is predicted that COVID-19 will maintain circulation alongside other seasonal or endemic respiratory viruses presenting with similar symptoms, such as influenza^8^. Multiplexed diagnostic screening for detection of SARS-CoV-2 variants and other respiratory pathogens must be made widely accessible to provide targeted treatments to patients and notify policy makers if more stringent measures are needed to control transmission.

Broad identification of virus variants and infectious pathogens can be achieved with genomic sequencing, but the necessary equipment and data processing required to conduct sequencing procedures is currently prohibitively expensive and complex for universal adoption^9^. Instead of sequencing, nucleic acid amplification tests (NAATs) typically used for infectious disease diagnosis may be readily modified to detect characteristic sequences of variants^10^. Although relatively easy to implement compared to sequencing, the increase in demand for NAATs during the SARS-CoV-2 pandemic has revealed severe deficiencies in public access to infectious disease diagnostics and aggravated existing shortages in testing capacity, supplies, and laboratory personnel^11^.

The gold standard NAATs for detection of SARS-CoV-2 RNA and other respiratory viruses use reverse-transcription polymerase chain reaction (RT-PCR)^12^. These tests provide the greatest sensitivity and specificity, but often require transport to high complexity laboratories in centralized test facilities which can lead to large backlogs with turnaround times of days or weeks^11,13^. Test results should ideally be delivered on-site at the testing location to facilitate recording of accurate surveillance data and to enable immediate notification of the test results to the patient for initiating quarantine or linkage to care. Currently available rapid NAAT platforms use expensive instruments and test cartridges making rapid screening for a large population with these systems unrealistic. Because of the long turnaround times and high cost per test of current NAAT platforms, many strategies for large scale testing opt for cheaper antigen tests^14^. Compared to PCR, these viral antigen tests have reduced sensitivity, higher rates of false-positives, and are not easily amenable to multiplexed detection of several genetic targets or pathogens^15,16^.

To address the need for affordable and accessible multiplexed NAATs with a fast turnaround time, we developed an imaging-based portable droplet magnetofluidic cartridge platform. Instead of traditional fluidic strategies for sample handling and assay automation, droplet magnetofluidic technologies use the movement of magnetic beads through discrete droplets of reagents to capture, purify, and transfer analytes for amplification and/or detection^17–20^. Recent developments of magnetofluidic cartridges have integrated static assay reagents isolated by immiscible liquid barriers into a low-cost plastic disposable^21– 23^. Magnetic transfer of beads along a hydrophobic surface within the cartridges obviates the need for precision fluidic channels and flow controllers that increase the complexity and cost of other automated NAAT platforms^24,25^. While previous studies using magnetofluidic cartridges were limited to a single reaction well per cartridge, this work achieves higher levels of multiplexed detection through a combination of duplexed PCR probe assays and a novel multi-elution aliquoting scheme to distribute nucleic acids.

Testing with our platform’s disposable cartridges enables sample-to-answer RT-PCR in under 30 minutes with detection of up to 4 genetic targets per test. Two different assay cartridges were designed – one for SARS-CoV-2 detection and differentiation of its variants, and another for multiplexed screening of SARS-CoV-2 with Influenza A, and Influenza B. We demonstrated the utility of our cartridge assays by testing with extracted RNA from clinical samples, nasopharyngeal swab eluates, and saliva samples. This platform presents a potent opportunity to expand access to screening SARS-CoV-2 variants and multiplexed respiratory pathogen testing in any setting with minimal training and immediate on-site reporting of results.

## Results

### Assay design and workflow

To conduct a test, the sample is first mixed with a buffer containing functionalized magnetic beads followed by loading the entire mixture into the sample port of the cartridge. Once sealed with an adhesive tab to prevent leakage of the sample, the cartridge is inserted into a slot in the side of the instrument (Fig. 1a, Supplementary Fig. 1). Identifying information for the sample is entered by the user using the instrument’s touchscreen interface followed by full automation of nucleic acid extraction, purification, and amplification by RT-PCR. The instrument conducts real-time analysis of fluorescent signals with fully interpreted results displayed on the screen in under 30 minutes. Each instrument has a compact footprint (14.5 cm ⨯ 21.6 cm ⨯ 14.5 cm) and built-in wireless connectivity for potential integration with laboratory information systems.

**Fig. 1.**
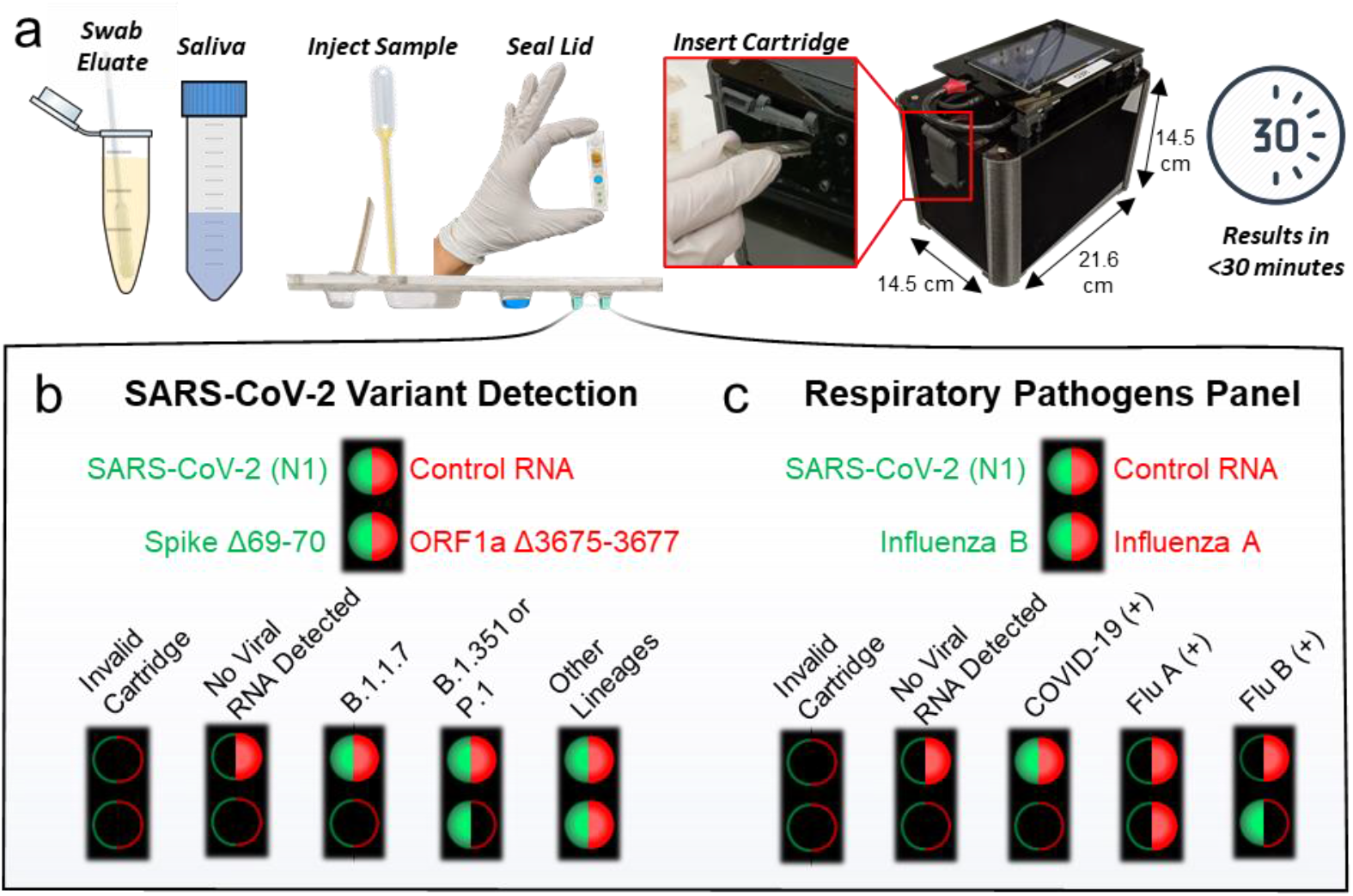
Cartridge platform operation. **a**, Nasal swab eluate or saliva is injected directly into the cartridge with magnetic beads followed by sealing the cartridge and inserting it into the instrument. After magnetofluidic sample preparation and PCR, the instrument reports the assay results on the built-in touchscreen within 30 minutes. **b**, Each PCR well contains two fluorescent probes in the FAM (green/left) or Cy5 (red/right) spectrum. Cartridges include a duplexed assay for the conserved N1 SARS-CoV-2 sequence and control RNA in the first well. The cartridge designed for detection of SARS-CoV-2 variants includes a duplexed PCR assay in the second well with probes spanning regions that contain deletions found in variants of concern. A lack of amplification in the second well indicates the presence of a mutation and can be used to classify the type of variant present. **c**, Cartridges designed for multiplexed detection of respiratory pathogens have a duplexed Influenza A and Influenza B PCR assay in the second well.

We designed two different cartridge assays (Fig. 1b-c) for either detection of SARS-CoV-2 variants of concern, or multiplexed diagnosis of SARS-CoV-2 with Influenza A and B. Both cartridges employ two duplexed PCR assays in separate wells containing hydrolysis probes labelled with FAM or Cy5/TYE fluorophores for a total of 4 target sequences per cartridge. To ensure cartridge reagents are functional and sample processing is fully completed, each cartridge assay detects a synthetic control RNA sequence^26^ that is pre-mixed in the magnetic bead solution. Detection of SARS-CoV-2 and control RNA are duplexed in the first well in both assays. A conserved nucleocapsid gene (N1) target sequence was adopted for universal detection of SARS-CoV-2^27^.

The cartridge for discrimination of SARS-CoV-2 variants uses the second PCR well for primers and probes designed by Vogel et al.^10^ to detect the presence of distinct mutations in the SARS-CoV-2 spike (Δ69-70) and ORF1a (Δ3675-3677) genes (Fig. 1b). The Δ69-70 mutation is associated uniquely with the B.1.1.7. variant that has shown high transmissibility.^4,5^ Meanwhile, the ORF1a deletion is found in B.1.1.7, B.1.351, and P.1 variants of concern ^10^. Therefore, if both the spike and the ORF1a mutation are present, the virus is classified as B.1.1.7. If the spike mutation is not detected, but the ORF1a mutation is, then the virus is classified as potentially B.1.351 or P.1. The PCR probes produce an amplification signal in virus lineages *not* included within the variants of concern, while signal dropout occurs if the described mutations are present. In the second cartridge designed for multiplexed detection of SARS-CoV-2 with influenza A and B, the second PCR well instead contains a duplex assay containing primers and probes for influenza A and B detection^28–30^ (Fig. 1c).

### Magnetofluidic cartridge design

The thermoplastic cartridge design in this work builds upon previous developments of magnetofluidic cartridges by incorporating greater flexibility in sample input volume, higher multiplexing of biomarkers, and more robust construction for easier handling^21–23^. Construction of the cartridges uses simple lamination techniques of three layers that have been laser-cut and thermoformed (Supplementary Fig. 2). All reagents are pre-loaded into extruded thermoformed wells of the cartridge (Fig. 2a) except for the magnetic beads which are mixed with the sample prior to loading into the cartridge. An immiscible layer of silicone oil provides an evaporation barrier and a fluidic interconnect between reagent wells for transfer of the magnetic beads. By isolating the reagents in thin-walled thermoformed wells, the thermal mass of the reaction can be spatially isolated for targeted, rapid thermocycling leading to faster turnaround times than traditional bulky PCR systems.

**Fig. 2.**
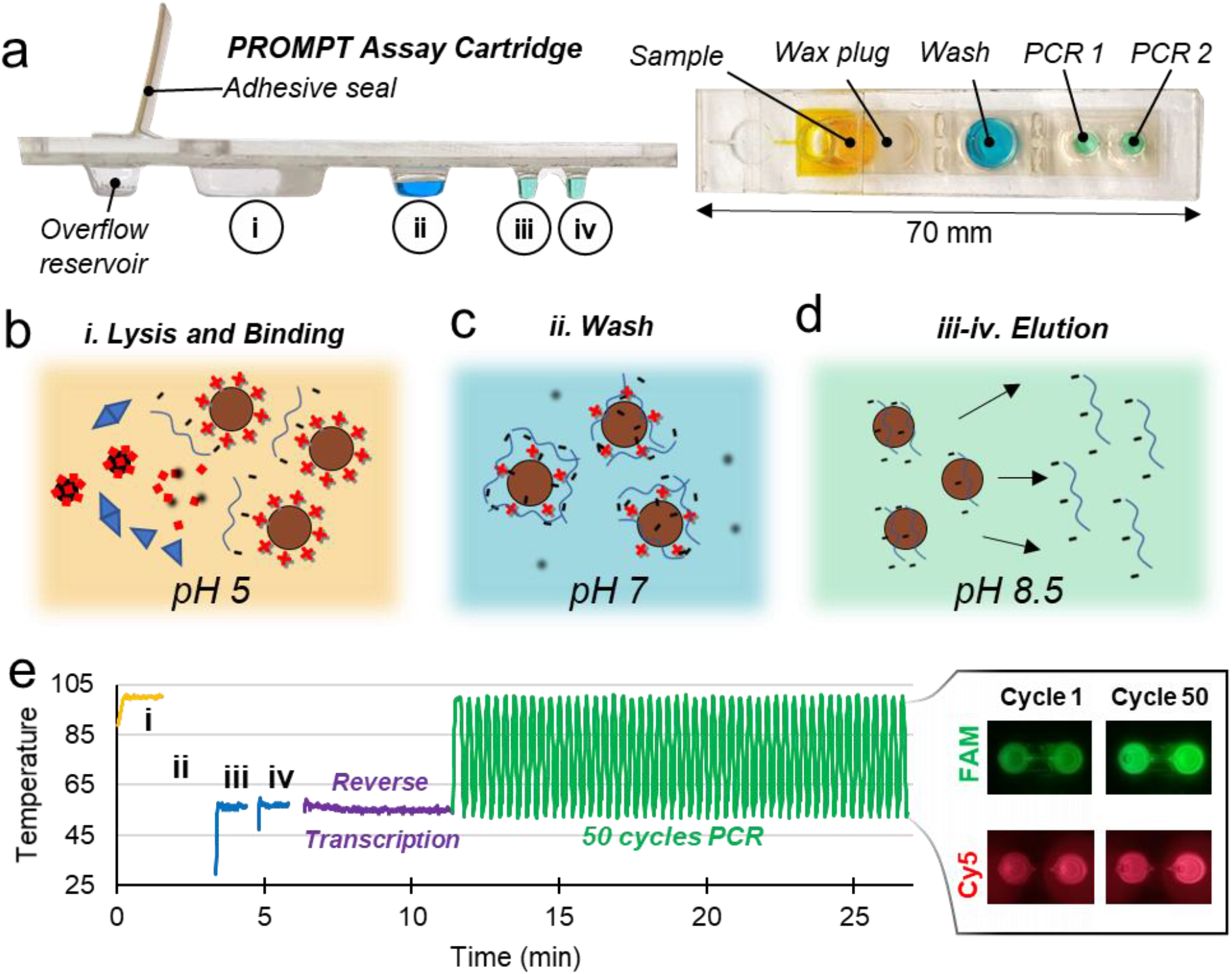
Cartridge design. **a**, The magnetofluidic cartridge contains preloaded assay reagents for sample purification and PCR. A layer of silicone oil fills the space within the cartridge between reagents, and a wax plug prevents the reagents from leaking during transport such that the first well remains empty for loading the sample. **b**, The first well is heated to 100°C for 80 seconds (i) to promote viral lysis for RNA capture and to release the wax plug at the base of the sample well to allow passage of magnetic beads. **c**, Bead transfer into the wash well (ii) promotes removal of salts, proteins, and other sample components that may inhibit PCR. **d**, Bead transfer into the PCR wells accompanied by heating to 55°C (iii, iv) allows sequential elution of the captured RNA. **e**, Plot of the temperature of previously described steps for sample preparation followed by one-pot reverse transcription and PCR with real-time 2-color fluorescence detection of both reaction wells at each cycle.

Once the sample is mixed with the magnetic bead buffer and loaded into the cartridge, the viral particles are lysed with surfactant and heating in the first well to allow for electrostatic binding of viral RNA to the charge-functionalized magnetic beads (Fig. 2b). Transfer of the beads into the second well exchanges the beads into a pH neutral wash buffer for removal of binding salts and any contaminants in the sample which may inhibit PCR (Fig. 2c). Finally, sequential transfer of the beads into the alkaline (pH 8.5) PCR wells allows for neutralization of the bead coating and partial release of captured RNA into each PCR reaction well (Fig. 2d) for amplification and fluorescence detection. To achieve under 30-minute turnaround time, sample preparation from lysis to completion of elution takes around 6 minutes, followed by 5 minutes of reverse transcription, and 50 cycles of PCR thermocycling in under 18 minutes (Fig. 2e).

The cartridge in this work includes several key innovations. A wax plug between the sample well and wash well seals off the oil and downstream reagents to immobilize all downstream fluids during transport and handling, which allows for full range of tilting and shaking the cartridge without reagent leakage. After the user injects the sample into the cartridge port, any excess sample can escape into an overflow reservoir and the port is sealed with an adhesive strip to provide an additional layer of safety from sample contamination and spill of infectious materials.

The most critical innovation in this work is the inclusion of an additional PCR well for higher levels of multiplexing coupled with a sequential elution strategy. Sequential elution takes advantage of the incomplete release of captured nucleic acids to aliquot RNA into separate reaction buffers. As the beads are exposed to each new buffer, the captured nucleic acids will be released until an equilibrium between the concentration of analyte on the bead surface and in the reaction buffer is reached. We have demonstrated this technique has potential to expand multiplexing up to at least six separate reactions (Supplementary Fig. 3). This flexibility in multiplexing provides an option to expand future cartridges to include additional targets for other SARS-CoV-2 variants or for a larger panel of pathogens.

### Instrument design and sample processing

The instrument contains all components necessary for transfer of magnetic beads through the cartridge, temperature control to melt wax seals and conduct RT-PCR, and optics for fluorescence excitation and detection (Fig. 3a-b). Instead of complex fluidics, valves, and pressure controllers typically found in microfluidic instrumentation, the components here include primarily low-cost hobby servo motors and off-the-shelf LED and CMOS camera parts (Supplementary Table 1). Once the cartridge is inserted into the instrument, it is detected with the CMOS camera, which uses the fluorescent outline of the PCR wells to determine if the cartridge is properly positioned. The fluorescence detection uses dual bandpass filters over the CMOS camera for emission, and over a 2-color LED for excitation to permit multi-color detection without moving parts by alternating blue and red LED illumination for FAM and Cy5/TYE fluorophores, respectively. Three servo motors automate application of the heat blocks to the cartridge and magnetic bead transfer (Supplementary Fig. 4). If the cartridge has been inserted fully, then the first servo motor involved rotates a shaft to mount both the sample heat block and PCR heat block onto the cartridge. With the heat blocks mounted, a power resistor heats the sample heat block to 100°C for 80 seconds to both promote viral lysis and melt the wax plug which then floats toward the top of the cartridge leaving a clear passage for transfer of the magnetic particles (Fig. 3c).

**Fig. 3.**
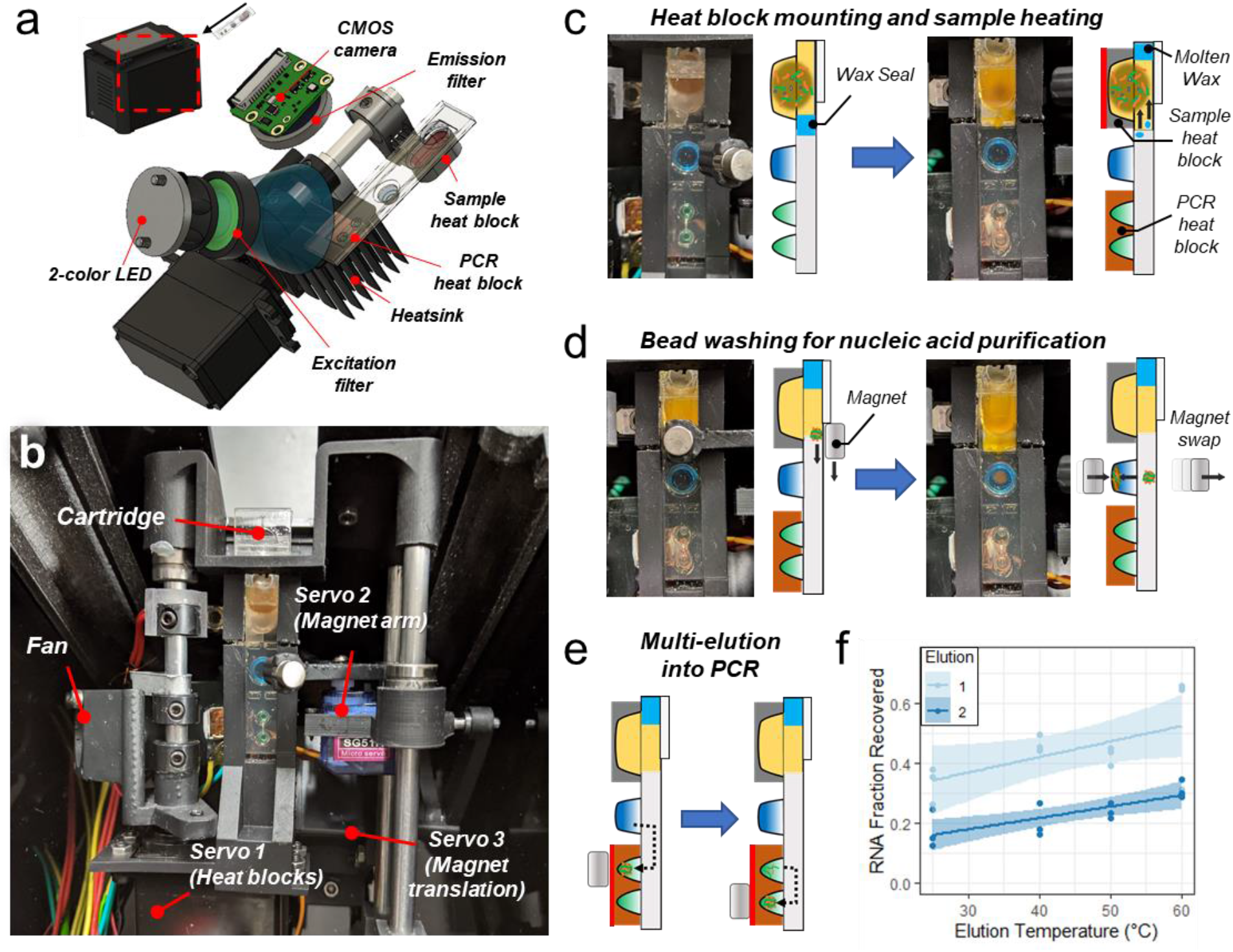
Instrumentation for automated sample preparation and multi-elution. **a**, Fluorescence detection optics and heat blocks assembly. **b**, Servo motor arrangement for (1) mounting heat blocks onto the cartridge, (2) swiveling magnets to the top and bottom of the cartridge for bead extraction and introduction into wells, and (3) translating the magnet arm along the cartridge for bead transfer between wells. **c**, Rotation of the heat blocks mounts them onto the cartridge followed by sample well heating to promote sample lysis and melt the wax seal. **d**, Translation of the top magnet from the sample well to the wash well followed by swiveling the magnet arm to swap the bottom magnet into close proximity with the cartridge pulls the beads into the wash buffer. **e**, Sequential transfer of beads into the first PCR well and then the second elutes captured RNA into both reactions. **f**, The first elution releases more RNA than the second elution with tunable release of the overall fraction of RNA by changing temperature of the buffer during elution. Both elution steps were run in triplicate at each temperature condition and shown here fit with a linear regression with 95% confidence interval bands.

A second servo motor swivels two opposing neodymium permanent magnets to the bottom or top of the cartridge to pellet beads into reagent wells or extract them into the oil layer. The third servo motor translates this magnet arm along the length of the cartridge for transfer of the beads between wells through the oil (Supplementary Video 1). This magnetic transfer paradigm allows the beads to be transferred anywhere along the long axis of the cartridge for compatibility with cartridge designs constructed with varying well number, dimensions, and positioning. With the wax melted, the beads are collected out of the sample well to the top of the cartridge and transferred into the wash buffer well to remove contaminants that might inhibit function of the downstream PCR assays (Fig. 3d). After alternate application of the top and bottom magnets for three repeated exchanges of the beads into and out of the wash buffer, the beads are finally transferred sequentially into the PCR wells (Fig. 3e). Each well receives the beads for 1 minute while the PCR buffers are heated to 55°C to facilitate release of the capture RNA and initiate reverse transcription. This multi-elution strategy permits some control over the release of RNA with higher elution temperature providing a higher fraction of RNA recovered in each well (Fig. 3f).

Immediately after elution, generation of cDNA and amplification is carried out with reverse transcription and PCR thermocycling. Temperature in both wells is simultaneously controlled by the PCR heat block with 2 second holds at 100°C for cDNA denaturation followed by 2 seconds at 55°C for annealing and extension. Miniaturization of the PCR heat block’s thermal mass and the use of copper’s high thermal conductivity (∼400 W/m-K) enables rapid changes in temperature powered by a heatsinked thermoelectric element (Supplementary Fig. 5). The optimized heat block has temperature ramp rates ranging between 6 to 10°C/s for both heating and cooling. When combined with time required for denaturation and annealing holds, controlled approaches to targeted temperatures, and image capture for fluorescence detection, this rapid thermocycling leads to completion of 50 cycles of PCR in less than 18 minutes.

### Assay cartridge analytical sensitivity and specificity

Throughout thermocycling, the CMOS camera takes a picture of the PCR wells for each fluorescence channel at the end of each cycle’s annealing step (Fig. 4a). The pixel intensity for each well is isolated and averaged to generate a real-time fluorescence curve (Fig. 4b), from which the cycle threshold (Ct) is determined with an automated thresholding algorithm (Supplementary Fig. 6). Detection of amplification and Ct calculation is conducted at the end of each cycle during the test for live reporting of results to the user. For high viral load samples (Ct <20), detection of targets may be reported within 18 minutes from insertion of the cartridge into the instrument.

**Fig 4.**
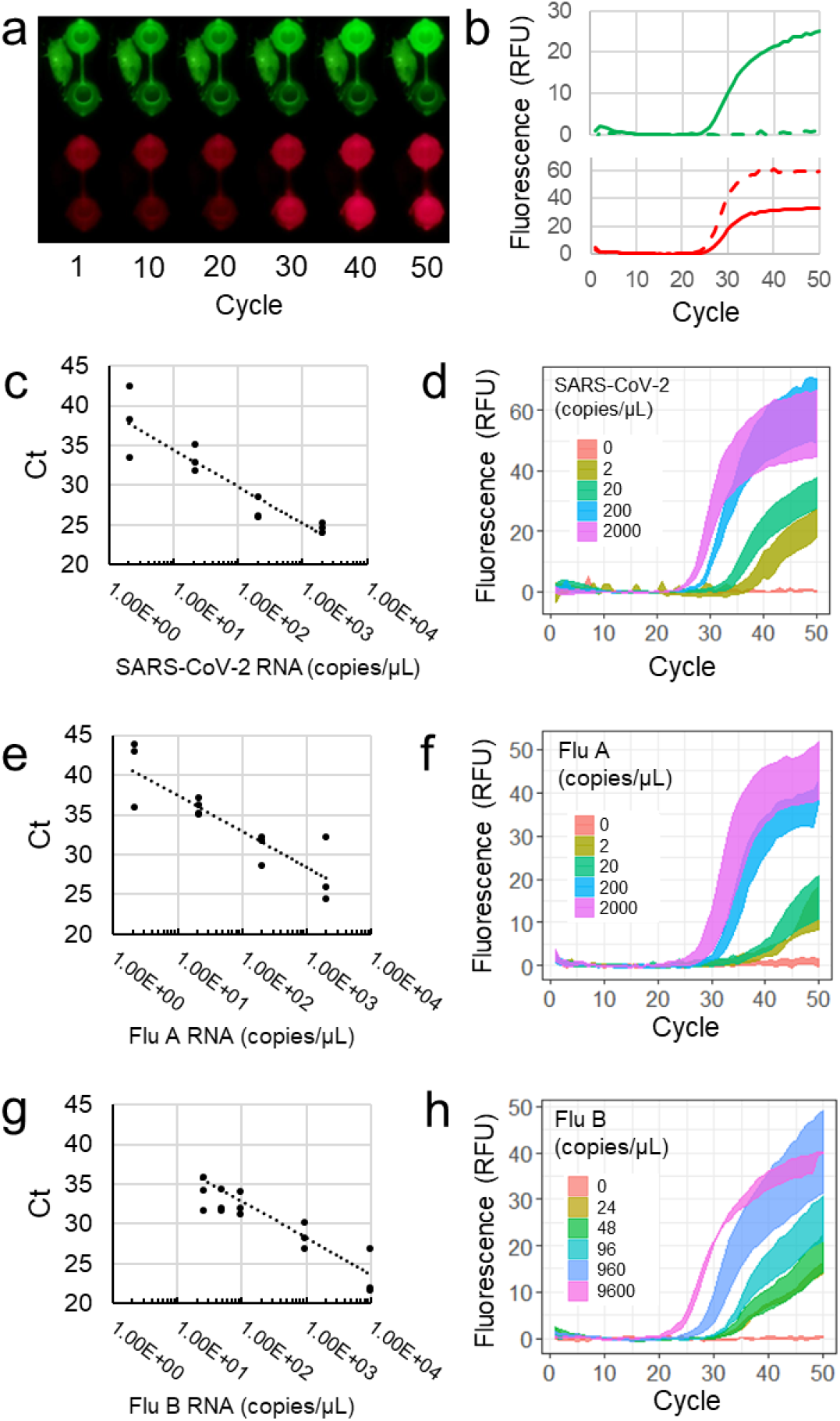
Cartridge PCR fluorescence curves and analytical sensitivity. **a**, Fluorescence images of PCR wells in the FAM and Cy5 channels taken at the annealing step for each cycle with corresponding real-time fluorescence curves plotted in (**b**). Solid lines and dotted lines in (**b**) correspond to the top and bottom wells respectively. Standard curves with Ct values and corresponding average of triplicate fluorescence curves with standard error are shown for SARS-CoV-2 (**c-d**), influenza A (**e-f**), and influenza B (**g-h**).

**Fig. 5.**
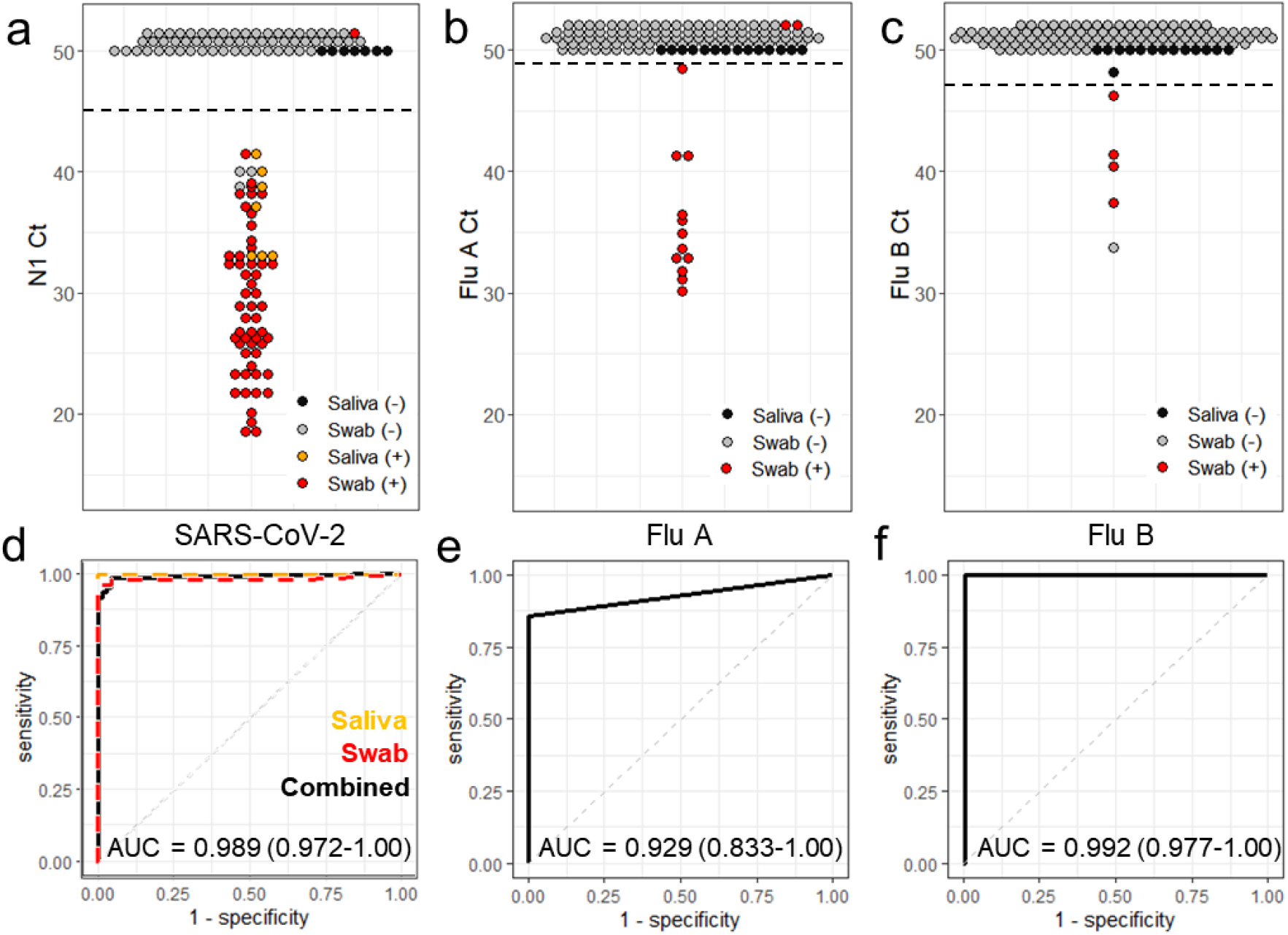
Clinical sample validation. **a-c**, PCR cycle threshold (Ct) values for swabs (*n* = 116) and saliva samples (*n* = 14) run in the multiplexed respiratory panel cartridges. Each point represents one sample with positive (+) and negative (-) classification determined by the benchtop comparator assay and denoted by color. All reactions with undetectable amplification are plotted with a Ct of 50 or higher. Cutoff Ct values for each assay are indicated with a horizontal dashed line. **d-f**, Corresponding receiver operator curves for the SARS-CoV-2, Flu A, and Flu B cartridge assays. Area under the curve (AUC) is indicated with 95% confidence interval.

Using the respiratory panel cartridge design, 50 µL samples containing serial dilutions of inactivated SARS-CoV-2, influenza A, or influenza B viral particles were loaded into cartridges with magnetic bead solutions. Each dilution was run in triplicate. Both SARS-CoV-2 and Influenza A were detectable in all replicates down to 2 copies/µL of sample (Figs. 4c-f), while Influenza B was detected down to 24 copies/µL of sample (Figs. 4g, h). We also demonstrated detection of SARS-CoV-2 spiked into saliva with a limit of detection of 12.5 copies/µL (Supplementary Fig. 7). To assess the specificity of the assay, the cartridges were run using a panel of 14 viral and bacterial pathogens (Supplementary Table 2). No cross-reactive false-positive amplification was detected for any pathogens in the specificity testing panel.

### Clinical sample validation

Using the cartridge for detection of SARS-CoV-2 variants we evaluated clinical samples from Johns Hopkins Hospital (JHH) as well as extracted RNA from B.1.1.7 and B.1.351 variants. All samples were previously classified by sequencing using the ARTIC protocol^31^ into three categories as either (1) B.1.1.7, (2) B.1.351/P.1, or (3) Other, indicating sample did not possess the characteristic mutations of current variants of concern (Table 1). All JHH samples (*n* = 4) amplified N1, ORF1a, and spike targets indicating they were not one of the variants of concern. Samples previously classified as B.1.1.7 variants (*n* = 4) by sequencing produced fluorescent amplification signals for N1 on cartridge, but no amplification of either the ORF1a or the spike targets and were accordingly classified properly. The samples characterized as B.1.351 (*n* = 3) by sequencing produced signals for N1 and spike, but did not amplify the ORF1a target resulting in proper classification as B.1.351 or P.1.

**Table 1.** Detection of SARS-CoV-2 Variants.

| Sample | N1-Ct | ORF1a-Ct | Spike-Ct | Classification |
| --- | --- | --- | --- | --- |
| JHH #1 | 30.39 | 32.57 | 32.64 | Other |
| JHH #2 | 30.78 | 34.44 | 33.85 | Other |
| JHH #3 | 28.73 | 38.10 | 27.49 | Other |
| JHH #4 | 28.33 | 45.57 | 27.54 | Other |
| B.1.1.7 #1 | 26.03 | Not detected | Not detected | B.1.1.7 |
| B.1.1.7 #2 | 32.03 | Not detected | Not detected | B.1.1.7 |
| B.1.1.7 #3 | 34.38 | Not detected | Not detected | B.1.1.7 |
| B.1.1.7 #4 | 30.13 | Not detected | Not detected | B.1.1.7 |
| B.1.351 #1 | 33.03 | Not detected | 37.59 | B.1.351 or P.1 |
| B.1.351 #2 | 28.29 | Not detected | 37.59 | B.1.351 or P.1 |
| B.1.351 #3 | 32.78 | Not detected | 31.49 | B.1.351 or P.1 |

To assess performance of the respiratory pathogen panel cartridges, we tested clinical swab eluates (*n* = 116) and passive drool saliva (*n* = 14). As a comparator assay, samples were first assessed with a benchtop assay adapted from the CDC recommended protocol (Supplementary Fig. 8)^32^. Of the swab samples, 54 were positive for SARS-CoV-2, 14 were Flu A positive, and 4 were Flu B positive using cutoff Ct values at 45, 49, and 47 respectively. Saliva samples contained 7 SARS-CoV-2 positives all of which were correctly classified with the cartridge assay. Only one of the SARS-CoV-2 samples went undetected in cartridge, which was a swab eluate with a relatively late Ct (37.2) by the benchtop comparator assay indicating a low viral titer. Of the negative SARS-CoV-2 swab samples, three false-positives were detected using cartridges, all with late Cts (> 39) indicative of possible low-level contamination from handling other positive samples. Sensitivity and specificity for detection of SARS-CoV-2 from swabs was 96.3% (95% confidence interval: 90.7-100%) and 95.2% (88.7-100%), respectively.

The cartridge Flu A assay missed detection of 2 out of the 14 positives which both had the lowest viral load of the Flu A samples (Ct >35), but yielded full concordance with negative samples, resulting in a sensitivity and specificity of 85.7 (64.3-100%) and 100%, respectively. For Flu B, all 4 positives were detected in cartridge with just one false-positive resulting in a specificity of 98.2% (95.5-100%). We have excluded 5 negative samples and 4 SARS-CoV-2 positive samples that produced invalid cartridge results with no amplification of the control or any other targets (Supplementary Table 3). These invalid results may be a consequence of samples with relatively high levels of inhibitory compounds.

## Discussion

There is an urgent need for affordable and mass-produced testing options that can provide rapid, multiplexed results for identification of variants and to allow future screening of SARS-CoV-2 with other diseases that produce similar respiratory symptoms. Compared to the intricate multi-component designs of current commercially available cartridges, our simple thermoplastic cartridges demonstrate great potential as a highly cost-effective and scalable solution, which is amenable to industrial manufacturing techniques such as roll-to-roll molding, lamination, and die-cutting.

Numerous *in vitro* diagnostic companies and researchers propose the use of isothermal NATs as PCR alternatives to leverage the sensitivity of RNA-based detection while reducing cost with simplified instrumentation or “instrumentation-free” testing^33,34^. These tests have been developed using isothermal techniques such as loop-mediated isothermal amplification (LAMP)^35,36^, recombinase polymerase amplification (RPA)^37^, or CRISPR-Cas based detection of viral RNA^38^. However, the need for manual processes to purify and concentrate sample RNA to achieve high sensitivity assays is often overlooked. Our magnetofluidic cartridges completely automate nucleic acid purification and concentration and minimize user intervention for reading results.

Our platform enables detection of multiple SARS-CoV-2 variants or other respiratory pathogens, while traditional rapid lateral-flow antigen tests and isothermal tests typically lack the ability to multiplex detection of several pathogens without either setting up multiple separate tests or requiring further post-processing steps for detection^39–41^. Post-processing of amplified products for readout of results adds an additional manual step which reduces the likelihood of adoption where a high-volume of tests requires minimal hands-on time. Furthermore, any handling of amplified product raises risks of contamination which would compromise test specificity.

Most low-cost rapid tests have minimal connectivity, making streamlined acquisition of patient results for surveillance difficult and can result in under-reporting of cases^42^. For widespread adoption, on-site multiplexed diagnostic methods need a user workflow as simple, fast, and affordable as lateral flow strips while maintaining connectivity for connecting to clinical databases for improving surveillance. This platform meets these needs in a compact user-friendly format that is compatible with various sample types including PBS, universal transport media, and saliva.

For realistic deployment, there are a few limitations to the current platform that must be addressed. In particular, the current cartridges are not shelf-stable for prolonged storage at room-temperature and are refrigerated or frozen prior to use. We are currently investigating techniques for built-in storage of dry reagents to allow stability at ambient conditions. To include testing for additional variants or respiratory pathogens, the cartridge would need to be expanded from the current 2-well design with additional PCR wells for higher multiplexing. Given the limited clinical sample volume available in this study, the assay design used a maximum 50 µL input per sample, though further improvement to sensitivity to prevent false-negatives may be achieved by adapting the cartridge and binding buffer to be compatible with larger volumes of sample.

## Methods

### RT-PCR assay composition

A 7.5-μL duplexed PCR probe assay was composed of 1X qScript XLT 1-Step RT-qPCR ToughMix (QuantaBio), 0.1 U/μL SpeedSTAR HS DNA polymerase (Takara Bio), 0.1 U/μL AccuStart II Taq DNA polymerase (QuantaBio), 1 mg/mL BSA (New England Biolabs), 0.1 % Tween-20 (Sigma Aldrich) and primer-probe pairs. For duplexed assay for N1 and control RNA, the assay contains 1 µM each N1 primer, 0.45 µM each Luciferase primer, 1 µM N1 probe and 0.25 µM Luciferase probe. For duplexed assay for influenza A and influenza B, the assay contains 0.5 µM each influenza A primer, 1 µM each influenza B primer, 0.25 µM influenza A probe and 0.5 µM influenza B probe. For duplexed assay of SARS-CoV-2 variant detection, the assay contains 0.67 µM each Yale Spike Δ69-70 primer, 0.3 µM each Yale ORF1a Δ3675-3677 primer, 0.2 µM Yale Spike Δ69-70 probe and 0.2 µM ORF1a Δ3675-3677 probe. All oligonucleotides, including primers and fluorescently labeled DNA probe (sequences in Supplementary Table 4) were purchased from Integrated DNA Technologies.

### Cartridge fabrication and assembly

The magnetofluidic cartridges were assembled from three thermoplastic layers (Supplementary Fig. 2). The bottom layer was fabricated by thermoforming 10 mil (∼0.25 mm) thick polyethylene terephthalate glycol (PETG) sheet (Welch Fluorocarbon) over 3D-printed molds (Form 2, Formlabs) designed in Fusion 360 (Autodesk) to produce extruded wells. The middle layer was laser-cut from 0.75 mm thick acrylic (ePlastics) with pressure-sensitive adhesive (PSA) (9472LE adhesive transfer tape, 3M) laminated on both sides. The top layer was laser-cut from 1.5 mm thick clear acrylic sheet (McMaster-Carr, USA) with Teflon tape (McMaster-Carr) laminated to one side and patterned by laser-etching.

To load reagents into the cartridge wells, the thermoformed section and acrylic middle layer were first joined with PSA, followed by dispensing 7.5 µL PCR solution and 50 µL wash buffer (W14, ChargeSwitch Total RNA Cell Kit, Invitrogen) into corresponding wells. With aqueous reagents pre-loaded, the cartridge was sealed by lamination with the top layer using the PSA on the other side of the middle layer. Once sealed, 420 µL silicone oil (100 cSt, Millipore-Sigma) was injected through the sample injection port to cover the wells and fill the remaining space within the cartridge except for the first well. To create the wax plug in the first well, 40 µL of molten docosane wax (Millipore-Sigma) was dispensed into the sample port and melted into the oil with a custom heating rig followed by cooling at room temperature to solidify. The cartridge was either used immediately or the sample injection port was sealed with adhesive tape (Scotch Magic Tape, 3M) and the cartridge stored on ice or frozen until use.

### Instrument design

Laser-cut and 3D-printed housing and fixtures of the instrument was designed in Fusion 360. External walls were laser-cut from 1/8” thick acrylic (McMaster-Carr) and 3D-printed components were fabricated using an SLA (Form 2, Black Resin, Formlabs) or FDM printer (Prusa Mini, Prusament PETG, Prusa research). A 5-inch HDMI touchscreen (Elecrow) was mounted on top of the instrument to allow user input with the graphic user interface (GUI) designed in python using the Tkinter library. Motorized actuation of an arm containing opposing neodymium magnets (K&J Magnetics) was implemented with a micro servo motor (TowerPro SG51R) mounted on a carriage guided along two aluminum rails by a second servo motor (Hitec HS-485HB). A third servo motor (Hitec HS-485HB) pivoted an aluminum rod to swivel the heat blocks onto the cartridge. The sample well heat block was custom machined out of 6061 aluminum and mounted onto a power resistor (Riedon PF1262-5RF1) with a steel M3 screw, while the PCR heat block was machined from 145 copper and mounted onto a thermoelectric element (Peltier Mini Module, Custom Thermoelectric) and heatsink using thermally conductive epoxy (Arctic Alumina Thermal Adhesive, Arctic Silver). Temperature of the heat blocks was monitored with a thermistor probe (GA100K6MCD1, TE Connectivity) epoxied directly adjacent to the wells. A 5V fan (Sunon) provided cooling to the heatsink.

Cartridges were illuminated using the red and blue channels of a 3-color RGB LED (Vollong) passed through a focusing lens (10356, Carclo) and dual bandpass excitation filter (59003m, Chroma). Fluorescence was captured with a CMOS camera (Pi NoIR Camera V2, Raspberry Pi) through a dual bandpass emission filter (535-700DBEM, Omega Optical). An Arduino Nano microcontroller coordinated control of the LEDs, fan, and motors, and a Raspberry Pi 3B+ ran the GUI, processed fluorescence images, monitored thermistor readings and provided current to the heat blocks via a motorshield (Dual TB9051FTG Motor Driver, Pololu). Power to the instrument was supplied with a 7.5V 45W wall adapter (MEAN WELL GST60A07-P1J).

### Cartridge limit of detection determination

Limit of detection of the cartridge assays was determined using contrived specimens of viral particles spiked into nasopharyngeal swab or saliva samples. Gamma-irradiated viral particles from SARS-Related Coronavirus 2 (Isolate USA-WA1/2020), Influenza A/Puerto Rico/8/1934-9VMC2(NR-29027), and Influenza B virus B/Nevada/03/2011 (BV) (NR-44023) were obtained through the BEI Resources Repository and stored at −80 °C upon receipt. To prepare the contrived samples, a serial dilution of viral particles were spiked into pooled (*n* = 4) clinical specimens (confirmed PCR-negative by the Johns Hopkins Clinical Microbiology Lab). Each concentration was tested a minimum of three times, and the limit of detection was determined when one of the replicates showed negative. Fifty microliters of swab eluate or five microliters of saliva was first mixed with 150 µL magnetic bead binding buffer consisting of 0.67 mg/mL ChargeSwitch beads, 0.5M KCl in 100mM aqueous MES, and 10^5^ copies of Luciferase RNA internal control (Promega). The entire mixture of sample and bead buffer was loaded into the sample port of the cartridge followed by insertion into the instrument for processing.

### Clinical sample testing

Clinical swab and saliva specimens were previously collected under Johns Hopkins IRB #00246027. Specimens were de-identified and blinded before testing. Nasopharyngeal swabs were eluted in 3 mL of Universal Transport Medium. Passive drooled saliva specimens were collected into an empty vessel. Five microliters of saliva was lysed with 50 µL of aqueous buffer containing 1% Triton X-100 and 1.2 units of Thermolabile Proteinase K (P8111S, New England Biolabs). 50 µL of swab eluate or 55 µL of saliva with lysis buffer was mixed with 150 µL magnetic bead binding buffer as previously described followed by loading the entire mixture into the sample port of the cartridge. The comparator assay for evaluation of clinical samples used a modified CDC testing protocol^32^ for swabs (Supplementary Methods, Supplementary Fig. 8) and FDA EUA authorized SalivaDirect protocol^43^ for saliva were employed to test all the clinical specimens on a Bio-Rad CFX96 Touch Real-Time PCR System as reference.

Testing of SARS-CoV-2 variants used RNA from clinical samples extracted using a chemagic 360 instrument (PerkinElmer) with clades of each sample previously characterized by sequencing^31^. 2 µL of extracted RNA was mixed with 150 µL magnetic bead binding buffer following by loading into the sample port of the cartridge.

## Data Availability

The main data supporting the results in this study are available within the paper and its supplementary information.

## Acknowledgements

We thank Emily Chang for her help with the instrument’s graphical user interface, and Shawna Lewis and Chun Huai (Alex) Luo for their assistance in collecting and preparing clinical samples. This work was supported by funding through the National Institutes of Health (R01AI138978, R01AI137272, R61AI154628).

## Supplementary Materials

### Supplementary Methods

#### Supplementary Figures List

**Fig. S1**. Loading Cartridges and Instrument Operation

**Fig. S2**. Cartridge Assembly

**Fig. S3**. Nucleic Acid Aliquoting with Sequential Elution

**Fig. S4**. Servo actuation for cartridge mounting and magnetic transfer

**Fig. S5**. Heat block simulation and design for rapid thermocycling

**Fig. S6**. Fluorescence processing algorithm

**Fig. S7**. Detection of SARS-CoV-2 in saliva

**Fig. S8**. Comparator assay evaluation

#### Supplementary Tables List

**Table 1**. Instrument bill of materials

**Table 2**. Specificity testing

**Table 3**. Clinical samples data

**Table 4**. PCR primers and probes

#### Supplementary Movies

**Movie S1**. On-cartridge sample processing

## References

1. Sutton, D., Fuchs, K., D’Alton, M. & Goffman, D. Universal Screening for SARS-CoV-2 in Women Admitted for Delivery. N. Engl. J. Med. 382, 2163–2164 (2020).

2. Kucharski, A. J. et al. Effectiveness of isolation, testing, contact tracing, and physical distancing on reducing transmission of SARS-CoV-2 in different settings: a mathematical modelling study. Lancet Infect. Dis. 20, 1151–1160 (2020).

3. Peto, J. et al. Weekly COVID-19 testing with household quarantine and contact tracing is feasible and would probably end the epidemic: Weekly Covid-19 testing. R. Soc. Open Sci. 7, 0–3 (2020).

4. Galloway, S. E. et al. Emergence of SARS-CoV-2 B.1.1.7 Lineage — United States, December 29, 2020–January 12, 2021. MMWR. Morb. Mortal. Wkly. Rep. 70, 95–99 (2021).

5. Volz, E. et al. Transmission of SARS-CoV-2 Lineage B.1.1.7 in England: Insights from linking epidemiological and genetic data. medRxiv 2020.12.30.20249034 (2021).

6. Lauring, A. S. & Hodcroft, E. B. Genetic Variants of SARS-CoV-2 - What Do They Mean? JAMA - J. Am. Med. Assoc. 325, 529–531 (2021).

7. Grubaugh, N. D., Hodcroft, E. B., Fauver, J. R., Phelan, A. L. & Cevik, M. Public health actions to control new SARS-CoV-2 variants. Cell 184, 1127–1132 (2021).

8. Veldhoen, M. & Simas, J. P. Endemic SARS-CoV-2 will maintain post-pandemic immunity. Nat. Rev. Immunol. (2021). doi:10.1038/s41577-020-00493-9

9. Gwinn, M., MacCannell, D. & Armstrong, G. L. Next-Generation Sequencing of Infectious Pathogens. JAMA 321, 893 (2019).

10. Vogels, C. B. et al. PCR assay to enhance global surveillance for SARS-CoV-2 variants of concern. medRxiv 351, 2021.01.28.21250486 (2021).

11. Babiker, A., Myers, C. W., Hill, C. E. & Guarner, J. SARS-CoV-2 Testing. Am. J. Clin. Pathol. 153, 706–708 (2020).

12. WHO. Laboratory testing for 2019 novel coronavirus (2019-nCoV) in suspected human cases. Interim guidance. (2020). Available at: https://www.who.int/publications/i/item/10665-331501. (Accessed: 6th December 2020)

13. McGarry, B. E., SteelFisher, G. K., Grabowski, D. C. & Barnett, M. L. COVID-19 Test Result Turnaround Time for Residents and Staff in US Nursing Homes. JAMA Intern. Med. 181, 556 (2021).

14. Peeling, R. W. et al. Serology testing in the COVID-19 pandemic response. Lancet Infect. Dis. 20, e245–e249 (2020).

15. Foundation for Innovative New Diagnostics (FIND). FIND evaluation update: SARS-CoV-2 Assays. Available at: https://www.finddx.org/covid-19/sarscov2-eval/. (Accessed: 6th December 2020)

16. Weissleder, R., Lee, H., Ko, J. & Pittet, M. J. COVID-19 diagnostics in context. Sci. Transl. Med. 12, 1–7 (2020).

17. Pipper, J. et al. Catching bird flu in a droplet. Nat. Med. 13, 1259–1263 (2007).

18. Shin, D. J. & Wang, T. H. Magnetic Droplet Manipulation Platforms for Nucleic Acid Detection at the Point of Care. Ann. Biomed. Eng. 42, 2289–2302 (2014).

19. Zhang, Y. & Nguyen, N.-T. Magnetic digital microfluidics – a review. Lab Chip 17, 994–1008 (2017).

20. Egatz-Gómez, A. et al. Discrete magnetic microfluidics. Appl. Phys. Lett. 89, (2006).

21. Shin, D. J. et al. Mobile nucleic acid amplification testing (mobiNAAT) for Chlamydia trachomatis screening in hospital emergency department settings. Sci. Rep. 7, 4495 (2017).

22. Shin, D. J., Trick, A. Y., Hsieh, Y. H., Thomas, D. L. & Wang, T. H. Sample-to-Answer Droplet Magnetofluidic Platform for Point-of-Care Hepatitis C Viral Load Quantitation. Sci. Rep. 8, (2018).

23. Trick, A. Y. et al. A portable magnetofluidic platform for detecting sexually transmitted infections and antimicrobial susceptibility. Sci. Transl. Med. 13, (2021).

24. Goldenberger, D. et al. Brief validation of the novel GeneXpert Xpress SARS-CoV-2 PCR assay. J. Virol. Methods 284, 113925 (2020).

25. Liotti, F. M. et al. Evaluating the newly developed BioFire COVID-19 test for SARS-CoV-2 molecular detection. Clin. Microbiol. Infect. 26, 1699–1700 (2020).

26. Johnson, D. R., Lee, P. K. H., Holmes, V. F. & Alvarez-Cohen, L. An internal reference technique for accurately quantifying specific mRNAs by real-time PCR with application to the tceA reductive dehalogenase gene. Appl. Environ. Microbiol. 71, 3866–3871 (2005).

27. Lu, X. et al. US CDC Real-Time Reverse Transcription PCR Panel for Detection of Severe Acute Respiratory Syndrome Coronavirus 2. Emerg. Infect. Dis. 26, 1654–1665 (2020).

28. Van Elden, L. J. R., Nijhuis, M., Schipper, P., Schuurman, R. & Van Loon, A. M. Simultaneous detection of influenza viruses A and B using real-time quantitative PCR. J. Clin. Microbiol. 39, 196–200 (2001).

29. Terrier, O. et al. Cellular transcriptional profiling in human lung epithelial cells infected by different subtypes of influenza A viruses reveals an overall down-regulation of the host p53 pathway. Virol. J. 8, 285 (2011).

30. World Health Organization. WHO information for the molecular detection of influenza viruses. (2020). Available at: https://www.who.int/influenza/gisrs_laboratory/Protocols_influenza_virus_detection_Jan_2020.pdf. (Accessed: 27th February 2021)

31. Thielen, P. M. et al. Genomic diversity of SARS-CoV-2 during early introduction into the Baltimore-Washington metropolitan area. JCI Insight 6, (2021).

32. Centers for Disease Control and Prevention (CDC). CDC 2019-Novel Coronavirus (2019-nCoV) Real-Time RT-PCR Diagnostic Panel, Revision: 06. (2020).

33. Peto, J., Hunter, D. J., Riboli, E. & Griffin, J. L. Unnecessary obstacles to COVID-19 mass testing. Lancet 396, 1633 (2020).

34. Woo, C. H., Jang, S., Shin, G., Jung, G. Y. & Lee, J. W. Sensitive fluorescence detection of SARS-CoV-2 RNA in clinical samples via one-pot isothermal ligation and transcription. Nat. Biomed. Eng. 4, 1168–1179 (2020).

35. Dao Thi, V. L. et al. A colorimetric RT-LAMP assay and LAMP-sequencing for detecting SARS-CoV-2 RNA in clinical samples. Sci. Transl. Med. 12, (2020).

36. Ganguli, A. et al. Rapid isothermal amplification and portable detection system for SARS-CoV-2. Proc. Natl. Acad. Sci. 117, 22727–22735 (2020).

37. Behrmann, O. et al. Rapid Detection of SARS-CoV-2 by Low Volume Real-Time Single Tube Reverse Transcription Recombinase Polymerase Amplification Using an Exo Probe with an Internally Linked Quencher (Exo-IQ). Clin. Chem. 66, 1047–1054 (2020).

38. Broughton, J. P. et al. CRISPR–Cas12-based detection of SARS-CoV-2. Nat. Biotechnol. 38, 870– 874 (2020).

39. Gootenberg, J. S. et al. Multiplexed and portable nucleic acid detection platform with Cas13, Cas12a and Csm6. Science (80-.). 360, 439–444 (2018).

40. Xu, G. et al. Paper-Origami-Based Multiplexed Malaria Diagnostics from Whole Blood. Angew. Chemie 128, 15476–15479 (2016).

41. Daher, R. K., Stewart, G., Boissinot, M. & Bergeron, M. G. Recombinase polymerase amplification for diagnostic applications. Clin. Chem. 62, 947–958 (2016).

42. Pradhan, R., Weber, L. & Recht, H. Lack of Antigen Test Reporting Leaves Country ‘Blind to the Pandemic’. Kaiser Health News (2020). Available at: https://khn.org/news/lack-of-antigen-test-reporting-leaves-country-blind-to-the-pandemic/. (Accessed: 21st April 2021)

43. FDA. Yale School of Public Health, Department of Epidemiology of Microbial Diseases SalivaDirect assay EUA Summary – Updated April 9, 2021. Available at: https://www.fda.gov/media/141192/download. (Accessed: 21st April 2021)

