## Supplementary Information for "Magnetofluidic platform for rapid multiplexed screening of SARS-CoV-2 variants and respiratory pathogens"

*Co-first author

**Co-corresponding author

### Supplementary Methods

#### *Modified CDC Protocol Comparator Assay*

The comparator assay protocol was modified from the CDC-recommended assay protocol[1]. Sample heating was omitted to simplify the procedure and polymerase concentration was increased to facilitate faster PCR thermal cycling. Briefly, one microliter of sample (swab eluate or saliva) was directly added to 9 µL of PCR reagent, which contain 1× master mix (qScript 1-Step Virus ToughMix, QuantaBio), 250 nM of each primer and TaqMan probe, 1% BSA, and 0.1% Tween-20. The mixture of sample and reagent was then incubated at 50°C for 10 minutes and 95°C for 2 minutes, followed by 50 cycles of thermal cycling from 95°C for 5 seconds to 60°C for 20 seconds. The assay was performed on a commercial real-time qPCR system BioRad CFX-96.

### Supplementary Figures

**Fig. S1.** Loading Cartridges and Instrument Operation

**Fig. S2.** Cartridge Assembly

**Fig. S3.** Nucleic Acid Aliquoting with Sequential Elution

**Fig. S4.** Servo actuation for cartridge mounting and magnetic transfer

**Fig. S5.** Heat block simulation and design for rapid thermocycling

**Fig. S6.** Fluorescence processing algorithm

**Fig. S7.** Detection of SARS-CoV-2 in saliva

**Fig. S8.** Comparator assay performance

### Supplementary Tables

**Table 1.** Instrument bill of materials

**Table 2.** Specificity testing

**Table 3.** Clinical samples data

**Table 4.** PCR primers and probes

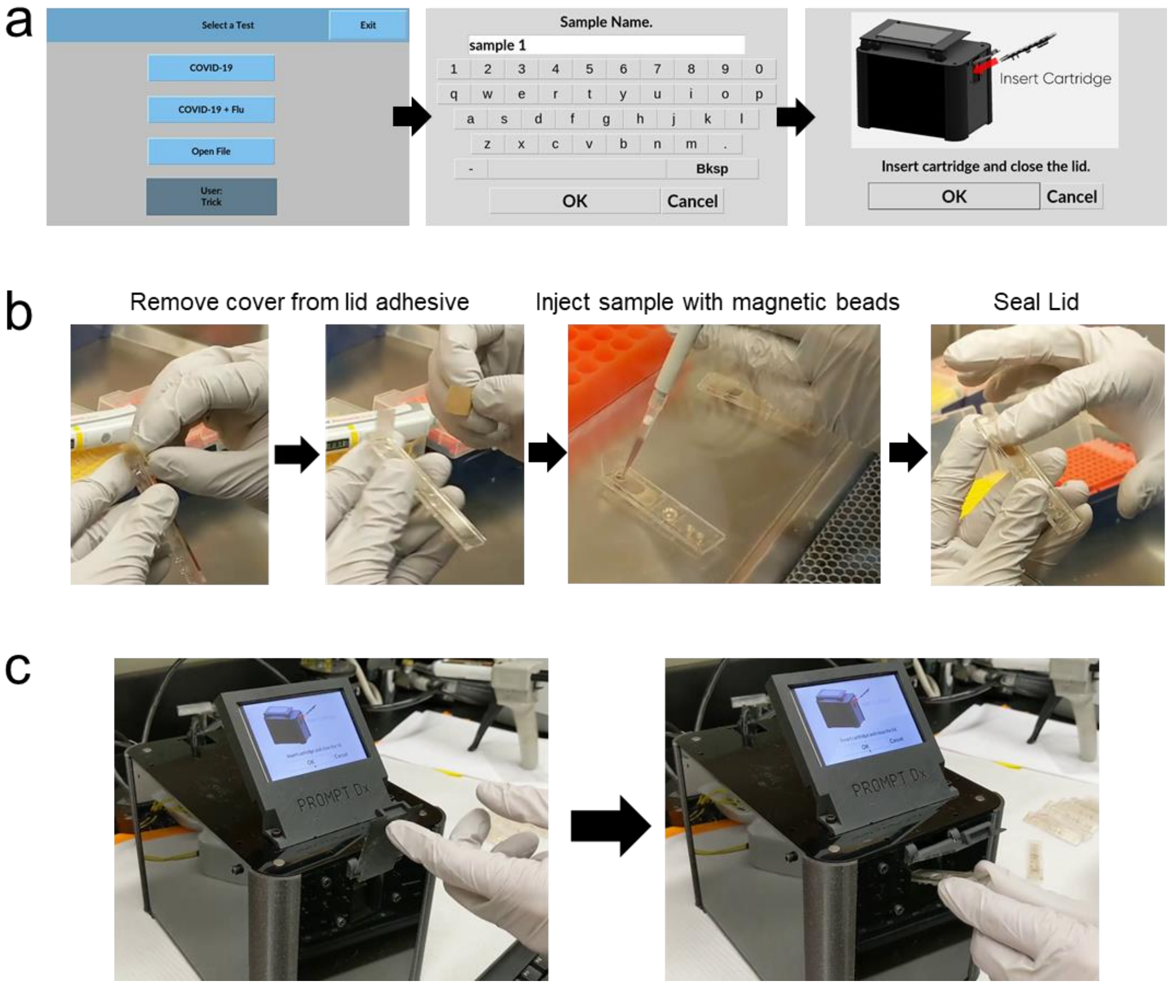


**Fig. S1.** Loading Cartridges and Instrument Operation. **a,** Graphic user interface for assay selection, entry of sample name, and confirmation of cartridge insertion into the instrument. **b,** To load the cartridge, first the paper cover is removed from the lid to expose a pressure-sensitive adhesive. Then the sample is mixed with magnetic beads and pipetted into the cartridge port followed by sealing the cartridge port with the adhesive lid. **c,** The cartridge is inserted into the instrument slot PCR wells first. Once the user presses “OK” on the touchscreen, the instrument verifies the proper positioning of the cartridge wells, mounts the heat blocks, and initiates sample preparation and RT-PCR.

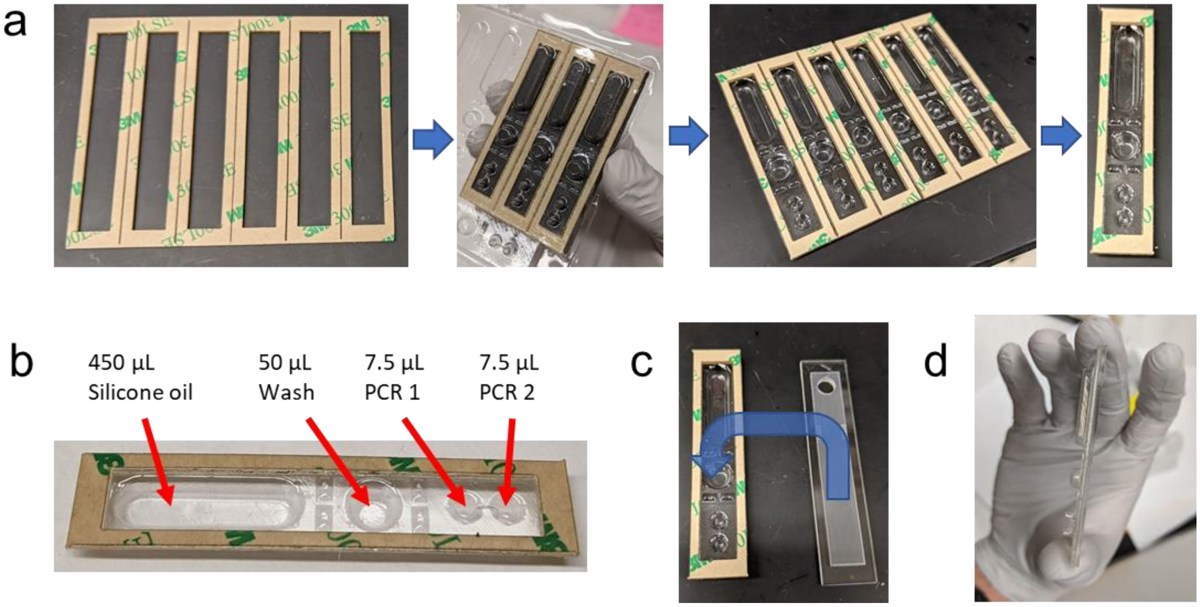


**Fig. S2.** Cartridge Assembly. **a,** The middle section of the cartridge is a laser-cut spacer acrylic with pressure sensitive adhesive laminated to both sides. One side of the adhesive cover is removed and 3D-printed alignment mold presses the spacer into the thermoformed wells. Each thermoform contains a row of 6 cartridges which are individually cut out prior to reagent loading. **b,** With the spacer section in place, silicone oil, wash buffer, and PCR buffers are dispensed directly into the thermoformed wells. **c,** A top layer with laser-patterned Teflon tape and a laser-cut sample port seals the cartridge using the top layer of pressure sensitive adhesive on the spacer. **d,** The cartridge is tilted to allow silicone oil to flow over the other reagent wells followed by dispensing of molten wax to plug the oil**.**

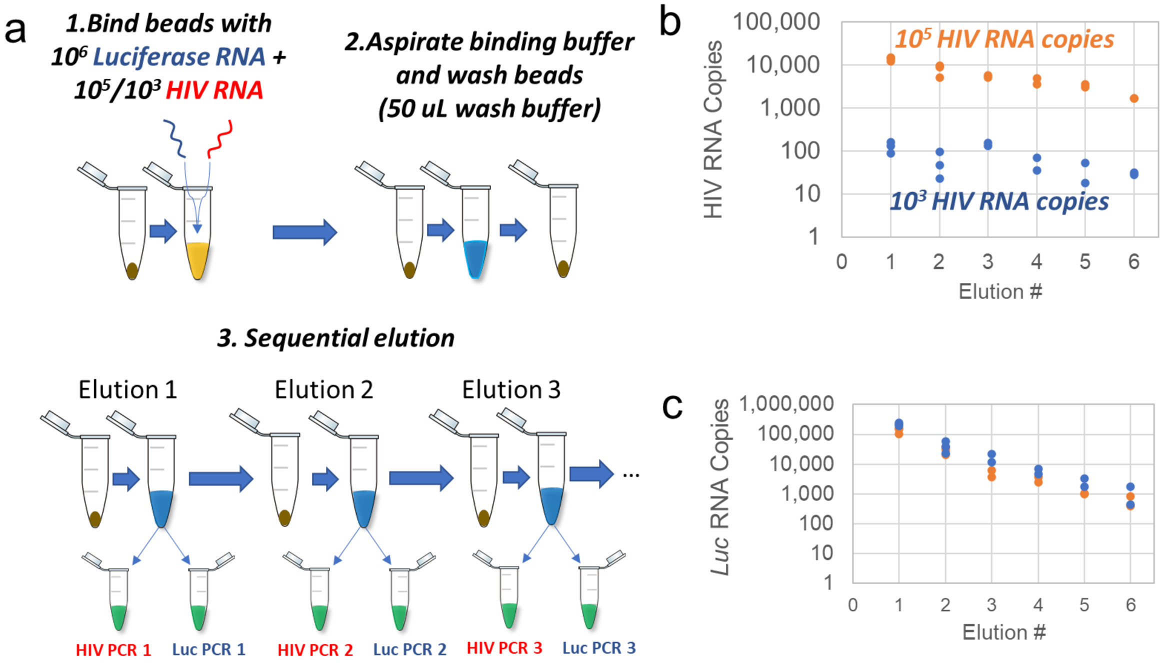


**Fig. S3.** Nucleic acid aliquoting with sequential elution. **a,** To evaluate higher levels of potential multiplexing through sequential elution, 4 µL ChargeSwitch beads were mixed with 10 µL ChargeSwitch binding buffer and 20 µL target containing $10^6$ copies of synthetic luciferase control RNA (L4561, Promega) with either $10^5$ or $10^3$ copies of synthetic HIV RNA (VR-3245SD ATCC). Beads were magnetically pelleted and the supernatant was aspirated and replaced with 50 µL ChargeSwitch wash buffer. After washing the beads, they were sequentially pelleted and resuspended into 10 µL ChargeSwitch elution buffer for a total of 6 elution steps. From each eluate, 1 µL was mixed into PCR assays for either amplification of HIV or luciferase (Luc) RNA. These assays contained 1X lyo-ready qPCR mix (Meridian Bioscience), 0.25 U PrimeScript RT enzyme (Takara Bio), 0.3 µM forward and reverse primers, and 0.25 µM probe (Table S4). Conditions for the first two elution steps were run in triplicate with duplicates for elution steps 3-6. **b,** HIV RNA recovery was calculated for each of the 6 elution steps for tubes containing both $10^5$ and $10^3$ copies as the initial input. HIV RNA was successfully detected in all eluates with a steady decreasing trend as the number of elutions increased. **c,** Luciferase RNA was also detected for all conditions with a trend that showed little deviation with either starting concentration of HIV target.

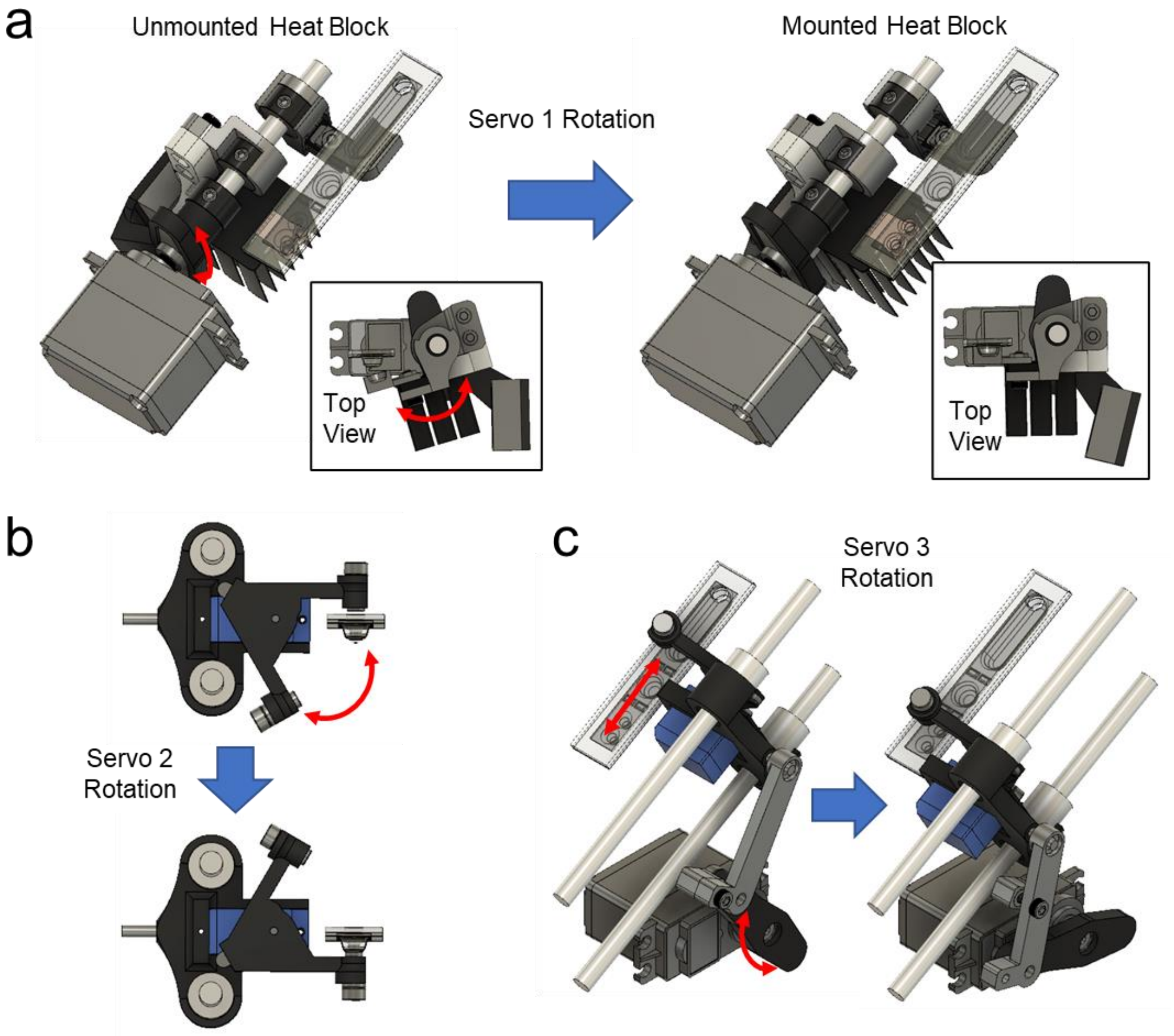


**Fig. S4.** Servo actuation for cartridge mounting and magnetic transfer**. a,** After insertion of the cartridge, the first servo rotates to mount the heat blocks onto the PCR and sample wells. **b,** Rotation of the second servo allows for transfer of magnetic beads into and out of cartridge wells. Here the rotation is shown pulling the beads from the planar PTFE-coated inner surface of the cartridge down into the reagent well. **c,** The third servo is connected to lever arms that actuate the second servo along a linear axis while the beads are captured on the PTFE inner surface to transfer the beads between wells.

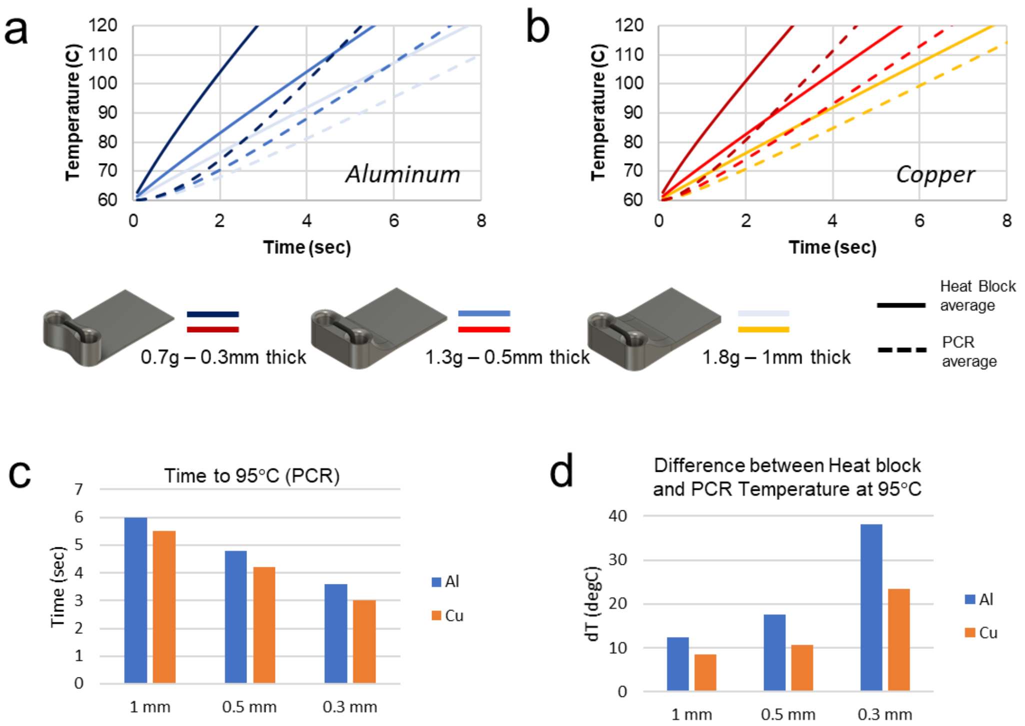


**Fig. S5.** Heat block simulation and design for rapid thermocycling. Transient heat transfer simulations were conducted in Solidworks 2019 using 3 different heat block models made from aluminum (**a**) or copper (**b**) with varying thickness. 7W constant heat power was supplied to the heat block in the rectangular section where the thermoelectric element would make contact. Average temperature for the heat block (solid lines) and 10 µL volumes of "PCR" (water) within each well (dashed lines) was tracked over 8 seconds. Reducing the thickness of the heat block resulted in faster heating, but larger differences between the heat block and PCR temperature. Copper's better thermal conductivity reduced the gradient between heat block temperature and PCR compared to aluminum. **c,** Comparison of time for the PCR to reach 95°C from 60°C starting temperature given the 7W constant heating in varying thickness copper (Cu) or aluminum (Al) heat blocks. **d,** Temperature difference between heat block and PCR when PCR first reaches 95°C indicates copper reduces the temperature gradient and requires less time to transfer heat from the thermoelectric to the PCR.

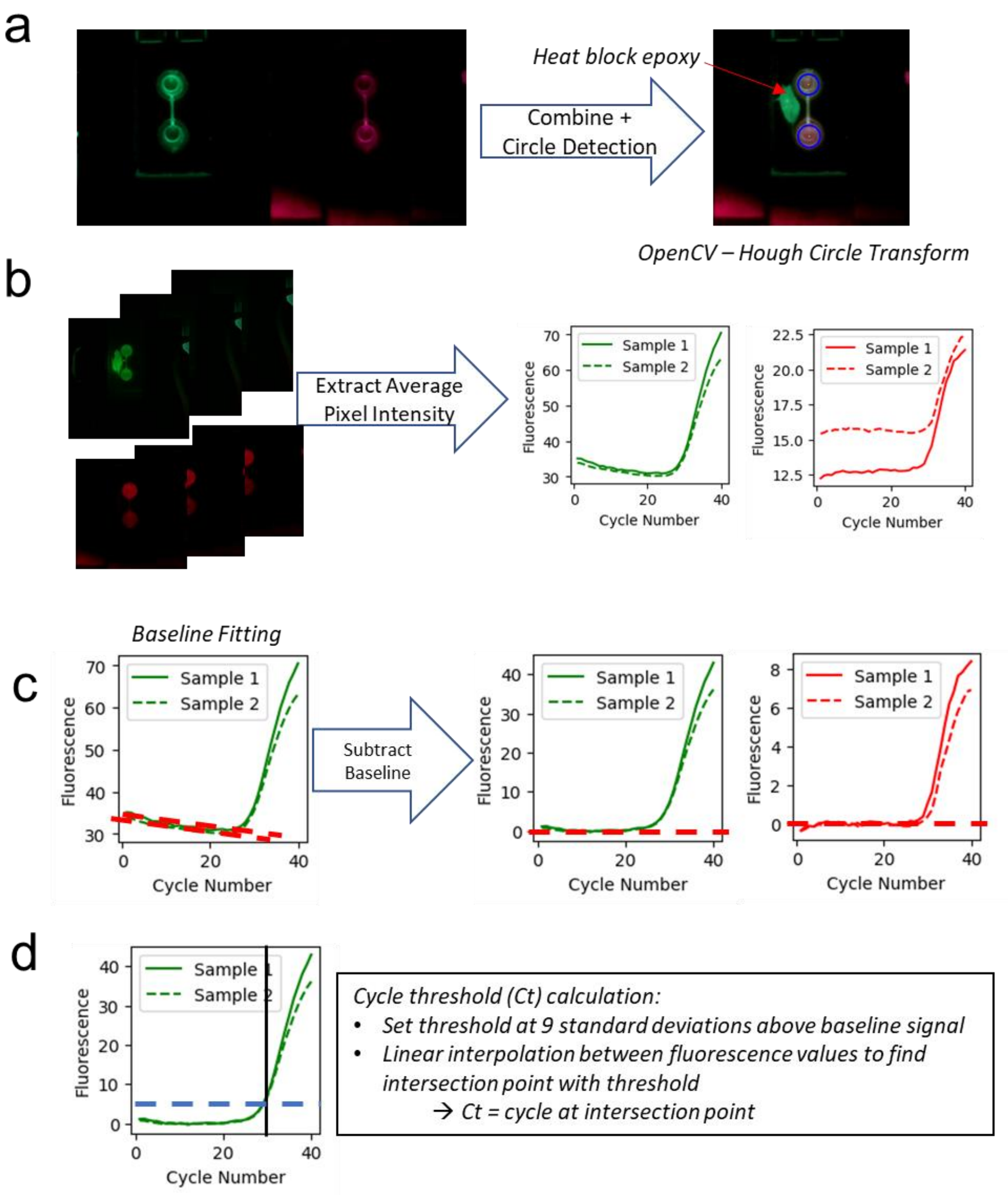


**Fig. S6.** Fluorescence processing algorithm. **a,** The cartridge is first imaged upon insertion to verify its position using the python OpenCV Hough Transform to detect circles. Images are taken in both the red and green channels and combined. If the circles are within a threshold distance of the calibrated cartridge position, then the heat block is mounted (evident by the fluorescent epoxy used to hold the thermistor to the heat block) and the circles are redetected after mounting. **b,** The detected circles are used to mask the images and extract the fluorescence for each individual well by averaging the pixel intensity. This results in the raw RT-PCR fluorescence curves. **c,** Fluorescence curves are baseline corrected by fitting the baseline with a linear regression curve and subtracting the resulting line from the whole set of points for each well and fluorescence channel. **d**, The baseline subtracted data is then used to calculate the cycle threshold (Ct) values for each curve using 9 standard deviations above the average fluorescence of the baseline as the threshold and interpolating the cycle and fluorescence data to find the cycle for the intersection point.

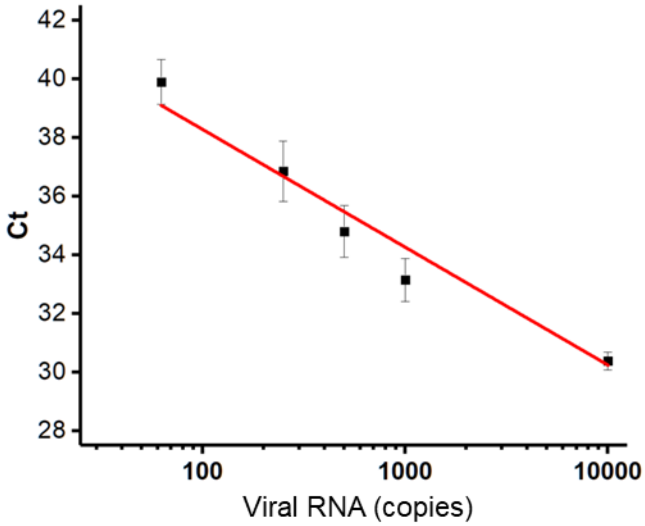


**Fig. S7.** Detection of SARS-CoV-2 in saliva. A serial dilution of known concentrations of gamma-irradiated viruses were spiked in pooled passive drool saliva samples (*n* = 4) collected from healthy donors. Five µL of each spiked saliva sample was mixed with 50 µL of magnetic bead buffer supplemented with thermo-labile proteinase K. The entire sample mix was then loaded into the cartridge for detection. The limit of detection was determined as 62.5 copies per 5 µL saliva input or 12.5 copies/µL, the lowest concentration of which at least three replicates were detected positive.

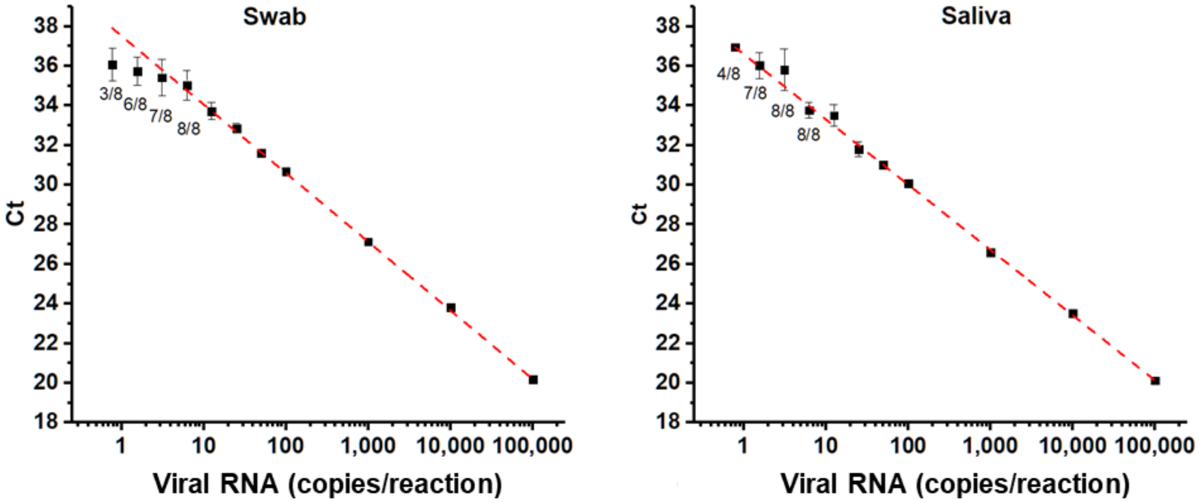


**Fig. S8.** Comparator assay evaluation. Using the modified CDC protocol, we were able to detect 3 copies/μL of g-irradiated SARS-CoVID-2 viruses in swab eluate and 1.5 copies/μL in saliva. The labels in the graphs indicate detected positive rate of eight replicates at each concentration. We defined the limit of detection as positive results for at least 7 out of 8 repeats.. Higher concentrations without number labels all tested positive (4 replicates). Red dashed lines represent linear regression.

### Supplementary Tables

**Table S1.** Instrument bill of materials

| Category | Part Description | Vendor | Model | Qty | Unit Price | Cost |
|---|---|---|---|---|---|---|
| Housing | M3 Screws | Amazon | DYWISHKEY 360 Pieces | 0.5 | $11.69 | $5.85 |
| Optics | Raspberry Picamera | Amazon | NoIR V2 8MP | 1 | $23.99 | $23.99 |
| Electronlcs | Arduino Nano (Elegoo) | Amazon | ELEGOO Nano pack of 3 | 0.333 | $19.98 | $6.65 |
| Electronlcs | Touchscreen | Amazon | ELECROW 800x480 Touch Screen | 1 | $49.99 | $49.99 |
| Electronlcs | HDMI Cables | Amazon | Cable Matters 3-pack | 0.33 | $12.99 | $4.33 |
| Electronlcs | USB Cables | Amazon | Sabrent 1ft - pack of 6 | 0.16 | $7.98 | $1.33 |
| Electronlcs | Power Supply Cord | Amazon | 6' | 1 | $7.42 | $7.42 |
| Electronlcs | 24 AWG stranded wire | Amazon | Silicone wire 24awg | 0.01 | $16.99 | $0.17 |
| Motors | Linear Ball Bearing | Amazon | LM6UU - 12 pieces | 0.25 | $7.99 | $2.00 |
| Motors | uxcell Ball Bearing | Amazon | ABEC-3 | 1 | $8.89 | $8.89 |
| Electronlcs | Raspberry Pi 3B+ | Canakit | 3B+ | 1 | $35.00 | $35.00 |
| Optics | Excitation Filter | Chroma | 59003m | 1 | $350 | $350.00 |
| Electronlcs | MCP3008 | Digikey | MCP3008-I/P | 1 | $2.19 | $2.19 |
| Motors | Sub Micro Servo | Digikey | SG51R | 1 | $5.95 | $5.95 |
| Temperature Control | 5 Ohm Power Resistor | Digikey | PF1262-5RF1 | 1 | $2.64 | $2.64 |
| Housing | 3D-printed Fixtures | Formlabs | Black Resin 1L | 0.2 | $149 | $29.80 |
| Electronlcs | Custom PCB | JLCPCB | Y6-2808702A | 0.2 | $19.55 | $3.91 |
| Magnets | Magnets - 3/16" x 1/4" | K&J Magnetics | D34-N52 | 2 | $0.47 | $0.94 |
| Magnets | Magnets - 1/4" x 1/8" | K&J Magnetics | D42-N52 | 6 | $0.43 | $2.58 |
| Electronlcs | BuckPuck | LuxeonStarLEDs | 3021-D-E-350 | 2 | $9.02 | $18.04 |
| Temperature Control | 25 mm heatsink | LuxeonStarLEDs | 25x25mm | 1 | $5.34 | $5.34 |
| Housing | Black Cast Acrylic Sheet | McMaster | 12"x24"x1/8" | 1 | $14.27 | $14.27 |
| Motors | 1/8" Aluminum Rod | McMaster | 3ft | 0.2 | $10.88 | $2.18 |
| Motors | 6 mm Aluminum Rod | McMaster | 3ft | 0.333 | $24.59 | $8.19 |
| Motors | 6mm set screw collar | McMaster | | 3 | $2.11 | $6.33 |
| Temperature Control | Easy-to-Machine 145 Copper Bar | McMaster | 1/4"x2"x1ft | 0.04 | $38.43 | $1.60 |
| Electronlcs | Power Supply 7.5V 6A | Mouser | MEAN WELL GST60A07-P1J | 1 | $19.30 | $19.30 |
| Temperature Control | 100kOhm Thermistor | Mouser | GA100K6MCD1 | 2 | $19.00 | $38.00 |
| Optics | Emission Filter | Omega | 535-700DBEM | 1 | $300 | $300.00 |
| Electronlcs | Dual TB9051FTG Motor Driver for Raspberry Pi | Pololu | 2761 | 1 | $21.95 | $21.95 |
| Electronlcs | 5V 5A Voltage Regulator | Pololu | 2851 | 1 | $14.95 | $14.95 |
| Motors | HS485HB Servo motors | RobotShop | HS-485HB | 2 | $17.99 | $35.98 |
| Optics | Vollong 3W RGB High Power LED | SuperBrightLEDs | VL-H01RGB00302 | 1 | $4.95 | $4.95 |
| Temperature Control | TEC | Custom Thermoelectric | 02301-9B30-32RU6A | 1 | $42.20 | $42.20 |
| | | | | | Total Cost: | $1,076.90 |

**Table S2.** Specificity testing

| Organism | Source | Concentration | Ct Value |
| --- | --- | --- | --- |
| Human coronavirus 229E | Bei Resources NR-52726 | 3.64× $10^{7}$ genome equivalents/mL | ND |
| Human coronavirus NL63 | Bei Resources NR-470 | 3.34× $10^{7}$ genome equivalents/mL | ND |
| Respiratory syncytial virus | Bei Resources NR-4052 | 136 ng/mL | ND |
| Human metapneumovirus | Bei Resources NR-22227 | 258 ng/mL | ND |
| *Acinetobacter baumannii* | ATCC 19606 | $10^{9}$ CFU/mL | ND |
| *Enterococcus faecalis* | ATCC 29212 | $10^{9}$ CFU/mL | ND |
| *Enterococcus faecium* | ATCC 35667 | $10^{9}$ CFU/mL | ND |
| *Escherichia coli* | ATCC 25922 | $10^{9}$ CFU/mL | ND |
| *Klebsiella pneumoniae* | ATCC BAA-1705 | $10^{9}$ CFU/mL | ND |
| *Morganella morganii* | ATCC 25830 | $10^{9}$ CFU/mL | ND |
| *Mycobacterium tuberculosis* | ATCC 25177 | $10^{9}$ CFU/mL | ND |
| *Staphylococcus aureus* | ATCC 29213 | $10^{9}$ CFU/mL | ND |
| *Streptococcus agalactiae* | ATCC 13813 | $10^{9}$ CFU/mL | ND |
| *Pseudomonas aeruginosa* | ATCC 27853 | $10^{9}$ CFU/mL | ND |

ND = no false-positive amplification detected

**Table S3.** Clinical samples date

| Sample ID | Comparator Assay/ Sample Type | Comparator Ct | Cartridge – N1 | Cartridge – Control | Cartridge – Flu B | Cartridge – Flu A |
|---|---|---|---|---|---|---|
| A10 | Flu A | 32.9 | 0.0 | 32.5 | 0.0 | 33.6 |
| A1 | Flu A | 31.5 | 0.0 | 38.6 | 0.0 | 36.5 |
| A2 | Flu A | 30.6 | 0.0 | 29.8 | 0.0 | 30.2 |
| A6 | Flu A | 29.3 | 0.0 | 34.5 | 0.0 | 31.1 |
| A5 | Flu A | 36.2 | 0.0 | 33.8 | 0.0 | 0.0 |
| A4 | Flu A | 30.3 | 0.0 | 35.6 | 0.0 | 32.7 |
| A12 | Flu A | 35.6 | 0.0 | 31.9 | 0.0 | 0.0 |
| A15 | Flu A | 29.8 | 0.0 | 34.0 | 0.0 | 34.9 |
| A11 | Flu A | 28.9 | 0.0 | 37.7 | 0.0 | 33.1 |
| A16 | Flu A | 32.8 | 0.0 | 0.0 | 0.0 | 41.2 |
| A17 | Flu A | 34.5 | 0.0 | 34.9 | 0.0 | 48.4 |
| A19 | Flu A | 29.7 | 0.0 | 31.3 | 0.0 | 35.9 |
| A20 | Flu A | 24.0 | 0.0 | 0.0 | 0.0 | 31.8 |
| A9 | Flu A | 32.6 | 39.0 | 0.0 | 0.0 | 41.4 |
| B14 | Flu B | 33.4 | 0.0 | 0.0 | 41.4 | 0.0 |
| B16 | Flu B | 34.6 | 0.0 | 34.3 | 40.4 | 0.0 |
| B20 | Flu B | 34.5 | 0.0 | 36.4 | 46.2 | 0.0 |
| B29 | Flu B | 27.5 | 0.0 | 0.0 | 37.4 | 0.0 |
| cv-neg21 | Negative Swab | 0.0 | 0.0 | 42.1 | 0.0 | 0.0 |
| cv-neg2-21 | Negative Swab | 0.0 | 0.0 | 32.5 | 0.0 | 0.0 |
| cv-neg2-22 | Negative Swab | 0.0 | 40.0 | 30.8 | 0.0 | 0.0 |
| cv-neg2-23 | Negative Swab | 0.0 | 0.0 | 31.2 | 0.0 | 0.0 |
| cv-neg2-24 | Negative Swab | 0.0 | 0.0 | 30.2 | 0.0 | 0.0 |
| cv-neg2-25 | Negative Swab | 0.0 | 0.0 | 34.4 | 0.0 | 0.0 |
| cv-neg2-26 | Negative Swab | 0.0 | 0.0 | 32.2 | 0.0 | 0.0 |
| cv-neg2-27 | Negative Swab | 0.0 | 0.0 | 28.4 | 0.0 | 0.0 |
| cv-neg2-28 | Negative Swab | 0.0 | 0.0 | 30.2 | 0.0 | 0.0 |
| cv-neg23 | Negative Swab | 0.0 | 0.0 | 29.7 | 0.0 | 0.0 |
| cv-neg24 | Negative Swab | 0.0 | 0.0 | 33.0 | 0.0 | 0.0 |
| cv-neg26 | Negative Swab | 0.0 | 0.0 | 31.6 | 0.0 | 0.0 |
| cv-neg28 | Negative Swab | 0.0 | 0.0 | 30.9 | 0.0 | 0.0 |
| cv-neg28 | Negative Swab | 0.0 | 0.0 | 27.8 | 0.0 | 0.0 |
| cv-neg29 | Negative Swab | 0.0 | 0.0 | 34.4 | 0.0 | 0.0 |
| cv-neg30 | Negative Swab | 0.0 | 0.0 | 31.6 | 0.0 | 0.0 |
| cv-neg37 | Negative Swab | 0.0 | 0.0 | 33.2 | 0.0 | 0.0 |
| cv-neg38 | Negative Swab | 0.0 | 0.0 | 29.2 | 0.0 | 0.0 |
| cv-neg39 | Negative Swab | 0.0 | 0.0 | 29.4 | 0.0 | 0.0 |
| cv-neg40 | Negative Swab | 0.0 | 0.0 | 40.5 | 0.0 | 0.0 |
| cv-neg41 | Negative Swab | 0.0 | 0.0 | 0.0 | 0.0 | 0.0 |
| cv-neg42 | Negative Swab | 0.0 | 0.0 | 0.0 | 0.0 | 0.0 |
| cv-neg43 | Negative Swab | 0.0 | 0.0 | 32.0 | 0.0 | 0.0 |
| cv-neg44 | Negative Swab | 0.0 | 0.0 | 32.9 | 0.0 | 0.0 |
| cv-neg45 | Negative Swab | 0.0 | 0.0 | 32.9 | 0.0 | 0.0 |
| cv-neg47 | Negative Swab | 0.0 | 0.0 | 32.9 | 0.0 | 0.0 |
| cv-neg48 | Negative Swab | 0.0 | 0.0 | 32.3 | 0.0 | 0.0 |
| cv-neg7 | Negative Swab | 0.0 | 0.0 | 34.1 | 0.0 | 0.0 |
| flu-neg1 | Negative Swab | 0.0 | 0.0 | 34.0 | 0.0 | 0.0 |
| flu-neg10 | Negative Swab | 0.0 | 0.0 | 48.3 | 0.0 | 0.0 |
| flu-neg13 | Negative Swab | 0.0 | 0.0 | 36.0 | 0.0 | 0.0 |
| flu-neg14 | Negative Swab | 0.0 | 0.0 | 28.7 | 0.0 | 0.0 |
| flu-neg15 | Negative Swab | 0.0 | 0.0 | 43.0 | 0.0 | 0.0 |
| flu-neg17 | Negative Swab | 0.0 | 0.0 | 32.1 | 0.0 | 0.0 |

| | | | | | | |
|---|---|---|---|---|---|---|
| flu-neg18 | Negative Swab | 0.0 | 0.0 | 33.5 | 0.0 | 0.0 |
| flu-neg19 | Negative Swab | 0.0 | 40.1 | 32.1 | 0.0 | 0.0 |
| flu-neg2 | Negative Swab | 0.0 | 0.0 | 33.8 | 0.0 | 0.0 |
| flu-neg20 | Negative Swab | 0.0 | 0.0 | 31.3 | 0.0 | 0.0 |
| flu-neg3 | Negative Swab | 0.0 | 0.0 | 35.2 | 0.0 | 0.0 |
| flu-neg5 | Negative Swab | 0.0 | 0.0 | 38.5 | 0.0 | 0.0 |
| flu-neg6 | Negative Swab | 0.0 | 0.0 | 32.6 | 0.0 | 0.0 |
| flu-neg8 | Negative Swab | 0.0 | 0.0 | 36.1 | 0.0 | 0.0 |
| saliva-201 | Saliva | 0.0 | 0.0 | 33.7 | 0.0 | 0.0 |
| saliva-203 | Saliva | 30.4 | 33.2 | 38.6 | 0.0 | 0.0 |
| saliva-204 | Saliva | 0.0 | 0.0 | 37.7 | 0.0 | 0.0 |
| saliva-205 | Saliva | 0.0 | 0.0 | 36.2 | 0.0 | 0.0 |
| saliva-206 | Saliva | 35.8 | 37.2 | 32.2 | 0.0 | 0.0 |
| saliva-207 | Saliva | 37.7 | 41.6 | 44.4 | 0.0 | 0.0 |
| saliva-208 | Saliva | 0.0 | 0.0 | 34.7 | 0.0 | 0.0 |
| saliva-209 | Saliva | 33.3 | 40.0 | 0.0 | 0.0 | 0.0 |
| saliva-210 | Saliva | 38.5 | 38.5 | 37.8 | 0.0 | 0.0 |
| saliva-211 | Saliva | 0.0 | 0.0 | 37.1 | 0.0 | 0.0 |
| saliva-212 | Saliva | 31.3 | 33.0 | 43.8 | 0.0 | 0.0 |
| saliva-213 | Saliva | 29.0 | 33.2 | 40.2 | 48.1 | 0.0 |
| saliva-NTC | Saliva | 0.0 | 0.0 | 35.9 | 0.0 | 0.0 |
| saliva-NTC | Saliva | 50.0 | 0.0 | 33.4 | 0.0 | 0.0 |
| 1 | SARS-CoV-2 | 23.9 | 25.0 | 46.2 | 0.0 | 0.0 |
| 2 | SARS-CoV-2 | 22.2 | 28.8 | 0.0 | 0.0 | 0.0 |
| 3 | SARS-CoV-2 | 26.3 | 23.6 | 0.0 | 0.0 | 0.0 |
| 4 | SARS-CoV-2 | 33.3 | 38.2 | 0.0 | 0.0 | 0.0 |
| 5 | SARS-CoV-2 | 26.1 | 26.1 | 0.0 | 0.0 | 0.0 |
| 6 | SARS-CoV-2 | 34.0 | 35.5 | 34.8 | 0.0 | 0.0 |
| 7 | SARS-CoV-2 | 0.0 | 0.0 | 0.0 | 0.0 | 0.0 |
| 8 | SARS-CoV-2 | 24.7 | 26.4 | 0.0 | 0.0 | 0.0 |
| 9 | SARS-CoV-2 | 32.2 | 32.2 | 0.0 | 0.0 | 0.0 |
| 10 | SARS-CoV-2 | 36.3 | 0.0 | 0.0 | 0.0 | 0.0 |
| 11 | SARS-CoV-2 | 37.2 | 0.0 | 33.4 | 0.0 | 0.0 |
| 12 | SARS-CoV-2 | 22.7 | 26.9 | 0.0 | 0.0 | 0.0 |
| 13 | SARS-CoV-2 | 24.3 | 30.0 | 0.0 | 0.0 | 0.0 |
| 14 | SARS-CoV-2 | 22.9 | 31.7 | 0.0 | 0.0 | 0.0 |
| 15 | SARS-CoV-2 | 33.3 | 30.7 | 32.8 | 0.0 | 0.0 |
| 16 | SARS-CoV-2 | 33.7 | 32.9 | 0.0 | 0.0 | 0.0 |
| Cov17 | SARS-CoV-2 | 34.2 | 37.1 | 30.7 | 0.0 | 0.0 |
| Cov18 | SARS-CoV-2 | 24.9 | 23.1 | 40.7 | 0.0 | 0.0 |
| Cov19 | SARS-CoV-2 | 30.2 | 32.2 | 32.2 | 0.0 | 0.0 |
| Cov20 | SARS-CoV-2 | 0.0 | 0.0 | 32.4 | 0.0 | 0.0 |
| Cov21 | SARS-CoV-2 | 20.7 | 21.9 | 48.0 | 0.0 | 0.0 |
| Cov22 | SARS-CoV-2 | 31.1 | 32.9 | 29.8 | 0.0 | 0.0 |
| Cov23 | SARS-CoV-2 | 30.0 | 39.1 | 0.0 | 0.0 | 0.0 |
| Cov24 | SARS-CoV-2 | 18.1 | 18.8 | 0.0 | 0.0 | 0.0 |
| Cov25 | SARS-CoV-2 | 25.9 | 26.7 | 30.7 | 0.0 | 0.0 |
| Cov26 | SARS-CoV-2 | 34.7 | 38.0 | 30.9 | 0.0 | 0.0 |
| Cov27 | SARS-CoV-2 | 18.4 | 23.6 | 0.0 | 0.0 | 0.0 |
| Cov28 | SARS-CoV-2 | 25.7 | 27.7 | 41.0 | 0.0 | 0.0 |
| Cov29 | SARS-CoV-2 | 32.8 | 32.2 | 29.2 | 0.0 | 0.0 |
| Cov30 | SARS-CoV-2 | 25.6 | 21.9 | 35.0 | 0.0 | 0.0 |
| Cov31 | SARS-CoV-2 | 27.5 | 26.0 | 0.0 | 0.0 | 0.0 |
| Cov32 | SARS-CoV-2 | 39.7 | 36.6 | 30.8 | 0.0 | 0.0 |
| Cov33 | SARS-CoV-2 | 0.0 | 0.0 | 31.5 | 0.0 | 0.0 |
| Cov34 | SARS-CoV-2 | 24.7 | 23.3 | 0.0 | 0.0 | 0.0 |
| Cov35 | SARS-CoV-2 | 18.3 | 18.4 | 0.0 | 0.0 | 0.0 |

| | | | | | | |
|---|---|---|---|---|---|---|
| Cov36 | SARS-CoV-2 | 25.4 | 24.9 | 0.0 | 0.0 | 0.0 |
| Cov37 | SARS-CoV-2 | 26.3 | 26.5 | 0.0 | 0.0 | 0.0 |
| Cov38 | SARS-CoV-2 | 26.5 | 25.7 | 33.0 | 0.0 | 0.0 |
| Cov39 | SARS-CoV-2 | 0.0 | 0.0 | 0.0 | 0.0 | 0.0 |
| Cov40 | SARS-CoV-2 | 28.5 | 26.9 | 37.0 | 0.0 | 0.0 |
| Cov41 | SARS-CoV-2 | 23.3 | 0.0 | 0.0 | 0.0 | 0.0 |
| Cov42 | SARS-CoV-2 | 28.8 | 28.2 | 0.0 | 0.0 | 0.0 |
| Cov43 | SARS-CoV-2 | 20.8 | 0.0 | 0.0 | 0.0 | 0.0 |
| Cov44 | SARS-CoV-2 | 22.0 | 21.9 | 0.0 | 0.0 | 0.0 |
| Cov45 | SARS-CoV-2 | 20.6 | 20.1 | 0.0 | 0.0 | 0.0 |
| Cov46 | SARS-CoV-2 | 29.0 | 26.5 | 0.0 | 0.0 | 0.0 |
| Cov47 | SARS-CoV-2 | 31.1 | 33.8 | 36.1 | 0.0 | 0.0 |
| Cov48 | SARS-CoV-2 | 24.2 | 25.5 | 47.0 | 0.0 | 0.0 |
| Cov49 | SARS-CoV-2 | 21.2 | 19.3 | 0.0 | 33.8 | 0.0 |
| Cov50 | SARS-CoV-2 | 33.5 | 32.6 | 38.0 | 0.0 | 0.0 |
| Cov51 | SARS-CoV-2 | 24.0 | 32.2 | 0.0 | 0.0 | 0.0 |
| Cov52 | SARS-CoV-2 | 19.7 | 38.4 | 36.1 | 0.0 | 0.0 |
| Cov53 | SARS-CoV-2 | 15.8 | 28.8 | 0.0 | 0.0 | 0.0 |
| Cov54 | SARS-CoV-2 | 0.0 | 0.0 | 39.0 | 0.0 | 0.0 |
| Cov55 | SARS-CoV-2 | 24.1 | 31.4 | 32.0 | 0.0 | 0.0 |
| Cov56 | SARS-CoV-2 | 25.6 | 21.6 | 0.0 | 0.0 | 0.0 |
| Cov58 | SARS-CoV-2 | 28.7 | 30.0 | 0.0 | 0.0 | 0.0 |
| Cov59 | SARS-CoV-2 | 26.9 | 41.4 | 0.0 | 0.0 | 0.0 |
| Cov60 | SARS-CoV-2 | 33.1 | 29.0 | 0.0 | 0.0 | 0.0 |
| Cov62 | SARS-CoV-2 | 30.0 | 24.0 | 0.0 | 0.0 | 0.0 |
| Cov63 | SARS-CoV-2 | 17.9 | 38.6 | 0.0 | 0.0 | 0.0 |
| Cov64 | SARS-CoV-2 | 0.0 | 0.0 | 0.0 | 0.0 | 0.0 |
| Cov65 | SARS-CoV-2 | 22.1 | 0.0 | 0.0 | 0.0 | 0.0 |
| Cov66 | SARS-CoV-2 | 34.3 | 34.3 | 40.2 | 0.0 | 0.0 |

**Ct of 0 indicates no amplification was detected. Orange cells indicate invalid cartridges excluded from analysis determined by absent amplification of all targets. Red cells indicate false-positive or false-negative as compared to the comparator assay.*

**Table S4.** PCR assay primers and probes

| Set name | Primer/probe | Sequence (5' → 3') | Reference |
|---|---|---|---|
| SARS-CoV-2 N1 | Forward primer | GACCCCAAAATCAGCGAAAT | 2 |
| | Reverse primer | TCTGGTTACTGCCAGTTGAATCTG | |
| | Probe | /FAM/ACCCCGCAT/ZEN/TACGTTTGGTGGACC/IABkF | |
| Control RNA | Forward primer | TACAACACCCCAACATCTTCGA | 3 |
| | Reverse primer | GGAAGTTCACCGGCGTCAT | |
| | Probe | /5TYE665/CGGGCGTGGCAGGTCTTCCC/3IAbRQSp/ | |
| Influenza A | Forward primer | CTTCTAACCGAGGTCGAAACGTA | 4 |
| | Reverse primer | GGTGACAGGATTGGTCTTGTCTTTA | |
| | Probe | /5TYE665/TCAGGCCCCCTCAAAGCCGAG/3IAbRQSp/ | |
| Influenza B | Forward primer | AAATACGGTGGATTAAACAAAAGCAA | 5 |
| | Reverse primer | CCAGCAATAGCTCCGAAGAAA | |
| | Probe | /56-FAM/CACCCATATTGGGCAATTTCCTATGGC/3IABkFQ | |
| Yale ORF1a Δ3675-3677 | Forward primer | TGCCTGCTAGTTGGGTGATG | 6 |
| | Reverse primer | TGCTGTCATAAGGATTAGTAACACT | |
| | Probe | /5Cy5/GTTTGTCTG/TAO/GTTTTAAGCTAAAAGACTGTG/3IAbRQSp | |
| Yale Spike Δ69-70 | Forward primer | TCAACTCAGGACTTGTTCTTACCT | |
| | Reverse primer | TGGTAGGACAGGGTTATCAAAC | |
| | Probe | /56-FAM/TTCCATGCT/ZEN/ATACATGTCTCTGGGA/3IABkFQ/ | |
| HIV | Forward primer | CATGTTTTCAGCATTATCAGAAGGA | 7 |
| | Reverse primer | TGCTTGATGTCCCCCCACT | |
| | Probe | /56-FAM/CCACCCCAC/ZEN/AAGATTTAAACACCATGCT AA/3IABkFQ/ | |

### Supplementary References


1. Centers for Disease Control and Prevention (CDC). CDC 2019-Novel Coronavirus (2019-nCoV) Real-Time RT-PCR Diagnostic Panel, Revision: 06. (2020).
2. Lu, X. *et al.* US CDC Real-Time Reverse Transcription PCR Panel for Detection of Severe Acute Respiratory Syndrome Coronavirus 2. *Emerg. Infect. Dis.* **26**, 1654–1665 (2020).
3. Johnson, D. R., Lee, P. K. H., Holmes, V. F. & Alvarez-Cohen, L. An internal reference technique for accurately quantifying specific mRNAs by real-time PCR with application to the tceA reductive dehalogenase gene. *Appl. Environ. Microbiol.* **71**, 3866–3871 (2005).
4. Terrier, O. *et al.* Cellular transcriptional profiling in human lung epithelial cells infected by different subtypes of influenza A viruses reveals an overall down-regulation of the host p53 pathway. *Virol. J.* **8**, 285 (2011).
5. Van Elden, L. J. R., Nijhuis, M., Schipper, P., Schuurman, R. & Van Loon, A. M. Simultaneous detection of influenza viruses A and B using real-time quantitative PCR. *J. Clin. Microbiol.* **39**, 196–200 (2001).
6. Vogels, C. B. *et al.* PCR assay to enhance global surveillance for SARS-CoV-2 variants of concern. *medRxiv* **351**, 2021.01.28.21250486 (2021).
7. Palmer, S. *et al.* New Real-Time Reverse Transcriptase-Initiated PCR Assay with Single-Copy Sensitivity for Human Immunodeficiency Virus Type 1 RNA in Plasma. *J. Clin. Microbiol.* **41**, 4531–4536 (2003).
